# Reports of myocarditis and pericarditis following mRNA COVID-19 vaccines: A systematic review of spontaneously reported data from the UK, Europe, and the US and of the literature

**DOI:** 10.1101/2021.09.09.21263342

**Authors:** Samantha Lane, Alison Yeomans, Saad Shakir

**Author notes:** Correspondence to: Miss Samantha Lane, Drug Safety Research Unit, Bursledon Hall, Blundell Lane, Southampton, Hampshire, SO31 1AA.

## Abstract

**Objectives:** To bring together spontaneously reported data from multiple countries to estimate reporting rate, and better understand risk factors for myocarditis and pericarditis following COVID-19 mRNA vaccines.

**Design:** Systematic review of spontaneously reported data from United Kingdom (UK), United States (US), and European Union/European Economic Area (EU/EEA) and of the literature.

**Data sources:** UK Yellow Card scheme, Vaccine Adverse Event Reporting System (VAERS), EudraVigilance were searched from date of vaccine launch to 14-16 March 2022. PubMed/MEDLINE and Embase were searched to 15 March 2022.

**Eligibility criteria:** We included publicly available spontaneous reporting data for “Myocarditis” and “Pericarditis” from UK, US, and EU/EEA following COVID-19 mRNA vaccines. Pharmacoepidemiological observational studies investigating myocarditis/pericarditis following mRNA COVID-19 vaccines were included (no restrictions on language or date). Critical Appraisal Skills Programme (CASP) tools assessed study quality.

**Data extraction and synthesis:** Two researchers extracted data. Spontaneously reported events of myocarditis and pericarditis were presented for each data source, stratified by vaccine, age, sex, and dose (where available). Reporting rates were calculated for myocarditis and pericarditis for each population. For published pharmacoepidemiological studies, design, participant characteristics, and study results were tabulated.

**Results:** Overall, 18,204 myocarditis and pericarditis events have been submitted to the UK, US, and EU/EEA regulators during the study period. Males represented 62.24% (n=11,331) of myocarditis and pericarditis reports. Most reports concerned vaccinees aged <40 years and were more frequent following a second dose. Reporting rates were consistent between the data sources. Thirty-two pharmacoepidemiological studies were included; results were consistent with our spontaneous report analyses.

**Conclusions:** Younger vaccinees more frequently report myocarditis and pericarditis following mRNA COVID-19 vaccines than older vaccinees. Results from published literature supported the results of our analyses.

**Strengths and Limitations of the Study:**

- This is the first study to bring together spontaneously reported data from the United Kingdom, United States, and Europe on myocarditis and pericarditis following mRNA COVID-19 vaccines.
- Results from this study provide evidence on the frequency of reported events of myocarditis and pericarditis following mRNA vaccines in different age groups, and by sex and vaccine dose; analyses of spontaneous reports were consolidated with results of published literature, identified by systematic review.
- Results may have been influenced by biases including different vaccination policies in each region examined, and publicity on events of myocarditis and pericarditis following mRNA vaccines.
- The study relied on outputs from spontaneous reporting systems in which the level of detail differed between the systems examined; furthermore, it is not possible to estimate incidence rates using spontaneous reports due to the lack of data on the exposed population, and there is no unvaccinated comparison group.
- There is an urgent need for further pharmacoepidemiological studies to be conducted to provide more accurate estimates of the frequency, clinical course, long term outcome, effects of treatment and impact on quality of life, to address many of the limitations of spontaneous reporting.

## 1.0 Introduction

Messenger RNA (mRNA) based vaccines have been extensively used world-wide in the fight against COVID19 that continues to pose a threat, with many countries initiating booster campaigns, yet these are the first in their class of vaccines to be approved for use, and as such continued monitoring of their safety is required. In the 15 months since first approval mRNA-based vaccines have had several adverse reactions documented, including myocarditis and pericarditis. Signals of myocarditis and pericarditis were first identified in Israel where there had been 148 cases of myocarditis reported within 30 days of vaccination, with the majority of these cases (n=121) reported after the second dose (1). Since the emergence of this signal, multiple countries have reported myocarditis and pericarditis following mRNA COVID-19 vaccines (2) and these events have been listed in the product information for both Pfizer-BioNTech (Comirnaty) and Moderna (Spikevax) mRNA COVID-19 vaccines (3-9). The identification of this safety signal early in the vaccination programme indicated that young males were at higher risk of developing myocarditis or pericarditis, particularly after the second dose of either mRNA based COVID-19 vaccine (2, 10-12). Myocarditis and pericarditis events following COVID-19 mRNA vaccines occur very rarely at a frequency of 10-20 events per 100,000 (13, 14), and the clinical course is typically mild with most cases making a full recovery (15).

Vaccination programmes around the world have differed in their roll-out and vaccine type used, with a general pattern that those most at risk of severe COVID-19 complications were prioritised for vaccination, followed by healthy adults and then children; in all these groups mRNA vaccines have been approved and used alongside adenovirus vector vaccines and inactivated virus vaccines around the world (4-9, 16, 17). Waves of COVID-19 infections in different countries also altered the speed of vaccine roll-out and the time interval between vaccine doses, thus interpretation of the data from several countries may reveal risk factors for myocarditis and pericarditis following exposure to COVID-19 mRNA vaccines. Here we collate spontaneous reports of myocarditis and pericarditis following COVID-19 vaccination, with a systematic review of the literature, to capture and interpret the evidence to date.

## 2.0 Materials and Methods

The data sources were spontaneous reporting system outputs of the UK Yellow Card scheme, the US Vaccine Adverse Event Reporting System (VAERS) via the CDC Wonder online tool, and the EU/EEA EudraVigilance system were used to estimate the frequency of reported cases of myocarditis and pericarditis following COVID-19 Vaccine Pfizer/BioNTech (Comirnaty) and COVID-19 Vaccine Moderna (Spikevax) (18-20). These systems collect unsolicited suspected adverse events to vaccines and medications from healthcare professionals and consumers. The process of spontaneous reporting requires suspected association with the mRNA vaccine to the event by the reporting individual. For reports following a mRNA COVID-19 vaccine, all reports coded “myocarditis” and “pericarditis” which had been spontaneously reported to these systems between the date of vaccine launch and the datalock point were counted. Cases were stratified by age, sex, and vaccine dose where these data were available.

The datalock point (defined as the date that searches were ran in the database) was 14 March 2022 for VAERS and EudraVigilance, and 16 March 2022 for the Yellow Card scheme. Data from the Yellow Card scheme is released weekly, with data up to 16 March 2022 the closest release to the 14 March 2022 datalock point used for VAERS and EudraVigilance.

The number of vaccinated individuals per vaccine brand in the UK, US, and EU/EEA were obtained from the websites of the MHRA, the CDC in the US, and the European Centre for Disease Prevention and Control (ECDC) up to the date closest to the datalock point for ADR spontaneous reports (11, 21, 22). Reporting rates of myocarditis and pericarditis per million vaccines administered were calculated for those who had received at least one dose of each vaccine brand.

### 2.1 Literature Review

A systematic literature review was conducted using PubMed/Medline and Embase literature databases. No review protocol was prepared.

The datalock point was 15 March 2022. Studies were included if they were observational in design (excluding case reports and case series), and involved at least one patient who experienced myocarditis, pericarditis, or myopericarditis following any mRNA COVID-19 vaccine (any case definition was accepted). Pre-print manuscripts were included if no peer-reviewed version was available. Studies of other designs and studies investigating other vaccines were excluded. Studies investigating cardiac effects of SARS-CoV-2 infection were excluded. No restrictions on language or date were applied.

The search terms used were:

(myocarditis OR pericarditis OR myopericarditis) AND (covid-19) AND (vaccine)

Data were extracted for study design, study period, vaccine of interest, population, number of cases of myocarditis or pericarditis, and where specified the percentage of cases who were male, age, and the vaccine dose after which the event occurred. Two reviewers extracted data. These data were tabulated.

Study quality was assessed by two reviewers, using the relevant Critical Appraisal Skills Programme (CASP) tool (available on request) (33). Each checklist covers a different study design and contains a series of questions to critically appraise the research.

### 2.2 Patient and Public Involvement

Patients and the public were not consulted during this study.

## 3.0 Results

### 3.1 Systematic review of the literature identified thirty-two observational studies

In order to collate information specific to address the issue identification of characteristics of myocarditis and pericarditis following COIVD-19 mRNA vaccines a detailed review of the literature was carried out, using the search terms specified in the materials of methods (Section 2.1). Initially 702 records were identified, following de-duplication, assessment for eligibility and quality 32 studies were included in our analysis (Figure 1). The information extracted in these studies has been assessed alongside the evidence from spontaneous reporting systems, below.

**Figure 1:**
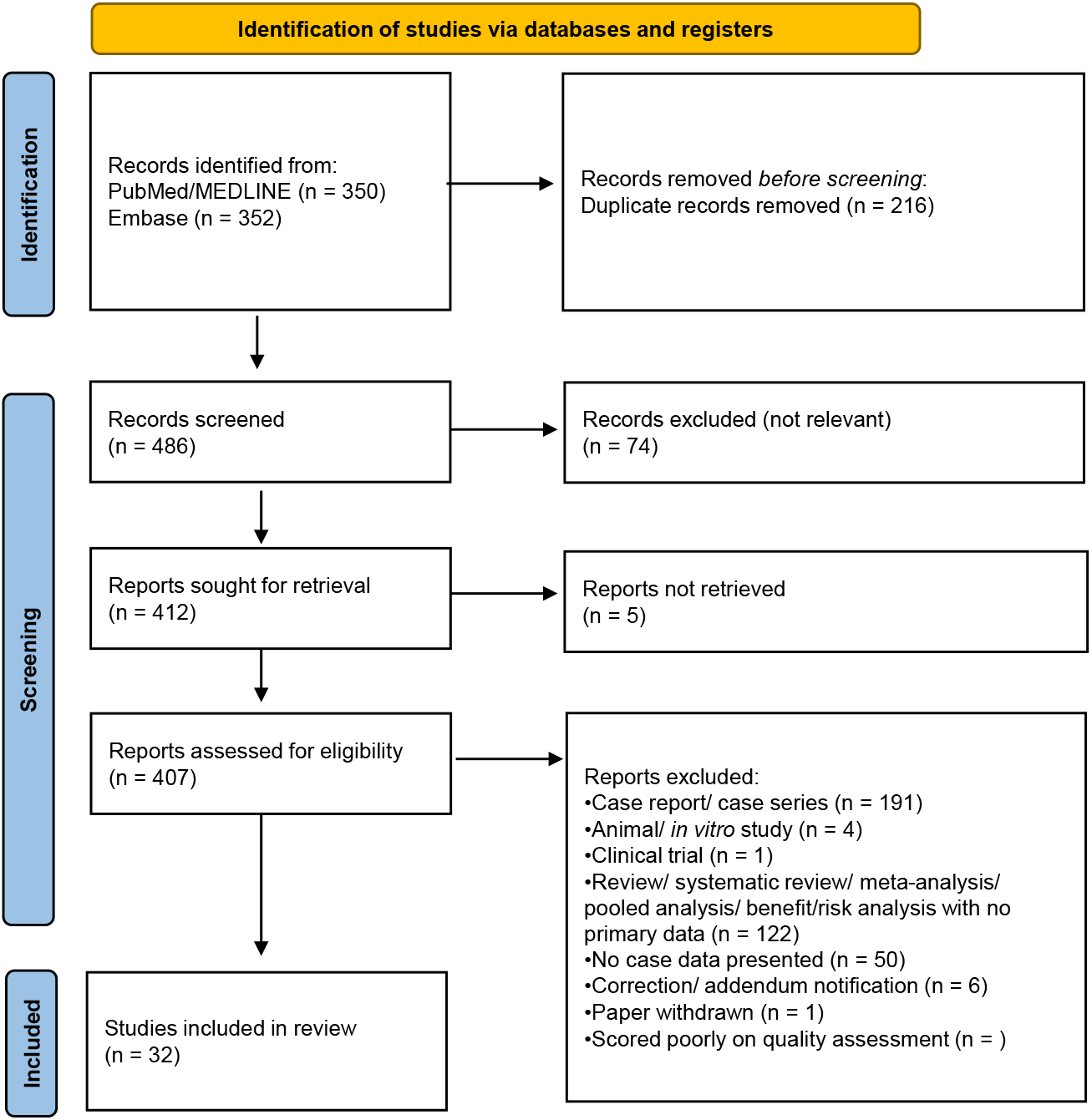
Flowchart detailing inclusion of studies for systematic review. Literature searches were carried out in PubMed/MEDLINE and simultaneously in Embase. The identified records were de-duplicated prior to screening to remove duplicate publications. Records were then screened to remove literature that did not meet the search criteria and eligibility was assessed prior to inclusion into the literature review.

### 3.2 Myocarditis and Pericarditis occur very rarely following COVID-19 mRNA vaccines

Overall, across the three spontaneous reporting databases examined covering the UK, US, and EU/EEA populations, there were a total of 18,204 events of myocarditis and pericarditis submitted to the regulators.

From the UK’s MHRA Yellow Card scheme, 1260 reports were following Comirnaty administration, and 325 reports were following Spikevax (Table 1; Figure 2A). As of 16 March 2022, it was estimated that 26.2 million first doses and 23.6 million second doses of Comirnaty had been administered in the UK (REF 3). Therefore, there were approximately 48.09 cases of myocarditis and pericarditis per million vaccinees who had received at least one dose of Comirnaty (Figure 2B). To the same date, approximately 1.6 million first doses and 1.5 million second doses of Spikevax had also been administered (REF 3). Of those who had received at least one dose of Spikevax in the UK, 203.13 cases of myocarditis and pericarditis had been reported per million vaccinated (Figure 2B).

**Table 1:** Reports of myocarditis and pericarditis submitted to Yellow Card scheme, by age and sex. Datalock point 16 March 2022.

|  | Comirnaty |  | Spikevax |  | Total |  |
| --- | --- | --- | --- | --- | --- | --- |
|  | n | % | n | % | n | % |
| **Age group (years)** |  |  |  |  |  |  |
| <18 Years | 70 | 5.68 | 0 | 0.00 | 70 | 4.50 |
| 18-29 Years | 372 | 30.19 | 114 | 35.19 | 486 | 31.23 |
| 30-39 Years | 299 | 24.27 | 92 | 28.40 | 391 | 25.13 |
| 40-49 Years | 137 | 11.12 | 48 | 14.81 | 185 | 11.89 |
| 50-59 Years | 92 | 7.47 | 22 | 6.79 | 114 | 7.33 |
| 60+ Years | 146 | 11.85 | 15 | 4.63 | 161 | 10.35 |
| Age not specified | 144 | 11.69 | 34 | 10.49 | 178 | 11.44 |
| **Sex** |  |  |  |  |  |  |
| Female | 492 | 39.94 | 109 | 33.64 | 601 | 38.62 |
| Male | 731 | 59.33 | 206 | 63.58 | 937 | 60.22 |
| Sex not specified | 37 | 3.00 | 10 | 3.09 | 47 | 3.02 |
| **Total** | **1260** | **100.00** | **325** | **100.00** | **1585** | **100.00** |

**Figure 2:**
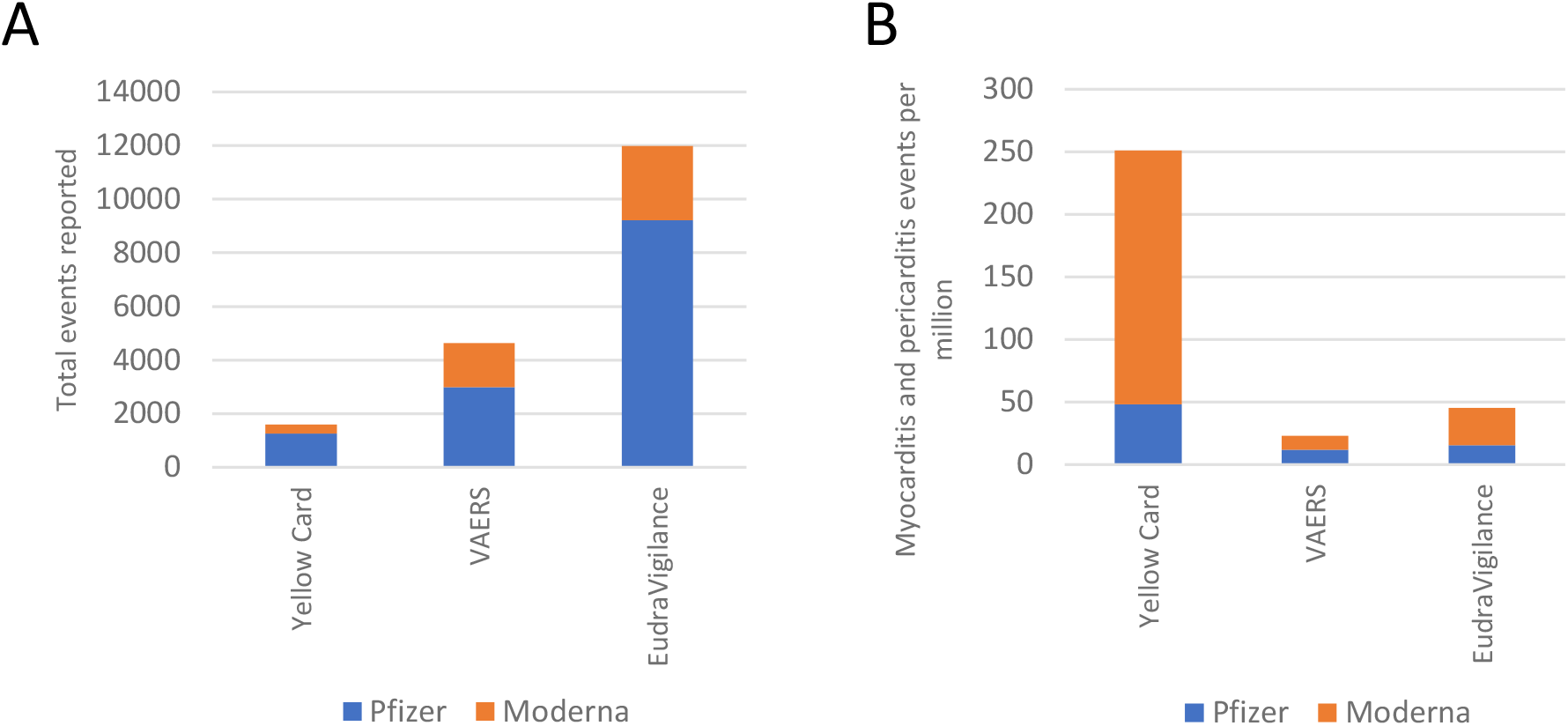
Event counts and reporting rates of myocarditis and pericarditis in the UK, US and EU. (A) Events of myocarditis and pericarditis reported to the Yellow Card scheme (UK), VAERS (US) and EudraVigilance (EU) reporting systems following any dose of COVID-19 mRNA vaccines separated by vaccine manufacturer combined to give total counts. (B) Reporting rates of myocarditis and pericarditis following any dose of COVID-19 mRNA vaccine separated by vaccine manufacturer and reporting region.

The US VAERS system contains 2986 reported events following Comirnaty and 1640 events following Spikevax (Table 2; Figure 2A). In the US, there had been 124.12 million vaccinees who had been fully vaccinated with Comirnaty (REF 29) giving a reporting rate of 14.70 cases of myocarditis and 9.36 cases of pericarditis per million fully vaccinated individuals, combined to 12.03 cases of myocarditis and pericarditis per million fully vaccinated with Comirnaty (Figure 2B). There had been 75.57 million people fully vaccinated with Spikevax (REF 29) therefore there were 12.35 cases of myocarditis reported per million fully vaccinated Spikevax recipients and 9.36 cases of pericarditis per million fully vaccinated Spikevax recipients, combined giving a reporting rate of both myocarditis and pericarditis as 10.86 per million Spikevax vaccinees (Figure 2B).

**Table 2:** Myocarditis and pericarditis events reported to VAERS overall. Datalock point 14 March 2022. Percentages per age group are presented.

| Age Group | Myocarditis |  |  |  |  |  |  |  | Pericarditis |  |  |  |  |  |  |  |
| --- | --- | --- | --- | --- | --- | --- | --- | --- | --- | --- | --- | --- | --- | --- | --- | --- |
|  | Male |  | Female |  | Sex Not Specified |  | Total |  | Male |  | Female |  | Sex Not Specified |  | Total |  |
|  | n | % | n | % | n | % | n | % | n | % | n | % | n | % | N | % |
| **Comirnaty (PfizerBioNTech)** |  |  |  |  |  |  |  |  |  |  |  |  |  |  |  |  |
| <6 months | 0 | 0.00 | 0 | 0.00 | 0 | 0.00 | 0 | 0.00 | 0 | 0.00 | 1 | 0.23 | 0 | 0.00 | 1 | 0.09 |
| 3-5 Years | 2 | 0.15 | 0 | 0.00 | 0 | 0.00 | 2 | 0.11 | 1 | 0.14 | 0 | 0.00 | 0 | 0.00 | 1 | 0.09 |
| 6-17 Years | 525 | 38.80 | 67 | 15.44 | 2 | 5.41 | 594 | 32.57 | 145 | 20.14 | 23 | 5.32 | 1 | 10.00 | 169 | 14.54 |
| 18-29 Years | 404 | 29.86 | 102 | 23.50 | 1 | 2.70 | 507 | 27.80 | 217 | 30.14 | 51 | 11.81 | 0 | 0.00 | 268 | 23.06 |
| 30-39 Years | 157 | 11.60 | 69 | 15.90 | 2 | 5.41 | 228 | 12.50 | 116 | 16.11 | 78 | 18.06 | 0 | 0.00 | 194 | 16.70 |
| 40-49 Years | 68 | 5.03 | 59 | 13.59 | 0 | 0.00 | 127 | 6.96 | 56 | 7.78 | 83 | 19.21 | 0 | 0.00 | 139 | 11.96 |
| 50-59 Years | 41 | 3.03 | 56 | 12.90 | 0 | 0.00 | 97 | 5.32 | 64 | 8.89 | 73 | 16.90 | 0 | 0.00 | 137 | 11.79 |
| 60-64 Years | 13 | 0.96 | 21 | 4.84 | 2 | 5.41 | 36 | 1.97 | 20 | 2.78 | 43 | 9.95 | 0 | 0.00 | 63 | 5.42 |
| 65-79 Years | 52 | 3.84 | 31 | 7.14 | 2 | 5.41 | 85 | 4.66 | 67 | 9.31 | 50 | 11.57 | 2 | 20.00 | 119 | 10.24 |
| 80+ Years | 5 | 0.37 | 4 | 0.92 | 0 | 0.00 | 9 | 0.49 | 12 | 1.67 | 9 | 2.08 | 0 | 0.00 | 21 | 1.81 |
| Not Specified | 86 | 6.36 | 25 | 5.76 | 28 | 75.68 | 139 | 7.62 | 22 | 3.06 | 21 | 4.86 | 7 | 70.00 | 50 | 4.30 |
| Total | 1353 | 100.00 | 434 | 100.00 | 37 | 100.00 | 1824 | 100.00 | 720 | 100.00 | 432 | 100.00 | 10 | 100.00 | 1162 | 100.00 |
| **Spikevax (Moderna)** |  |  |  |  |  |  |  |  |  |  |  |  |  |  |  |  |
| <6 months | 0 | 0.00 | 0 | 0.00 | 0 | 0.00 | 0 | 0.00 | 0 | 0.00 | 0 | 0.00 | 0 | 0.00 | 0 | 0.00 |
| 3-5 Years | 0 | 0.00 | 0 | 0.00 | 0 | 0.00 | 0 | 0.00 | 0 | 0.00 | 0 | 0.00 | 0 | 0.00 | 0 | 0.00 |
| 6-17 Years | 5 | 0.78 | 1 | 0.37 | 2 | 10.53 | 8 | 0.86 | 1 | 0.26 | 0 | 0.00 | 0 | 0.00 | 1 | 0.14 |
| 18-29 Years | 317 | 49.15 | 64 | 23.79 | 1 | 5.26 | 382 | 40.94 | 130 | 33.33 | 62 | 20.39 | 0 | 0.00 | 192 | 27.16 |
| 30-39 Years | 135 | 20.93 | 51 | 18.96 | 0 | 0.00 | 186 | 19.94 | 59 | 15.13 | 38 | 12.50 | 1 | 7.69 | 98 | 13.86 |
| 40-49 Years | 62 | 9.61 | 51 | 18.96 | 0 | 0.00 | 113 | 12.11 | 57 | 14.62 | 54 | 17.76 | 0 | 0.00 | 111 | 15.70 |
| 50-59 Years | 41 | 6.36 | 37 | 13.75 | 1 | 5.26 | 79 | 8.47 | 52 | 13.33 | 52 | 17.11 | 0 | 0.00 | 104 | 14.71 |
| 60-64 Years | 15 | 2.33 | 24 | 8.92 | 0 | 0.00 | 39 | 4.18 | 24 | 6.15 | 31 | 10.20 | 1 | 7.69 | 56 | 7.92 |
| 65-79 Years | 29 | 4.50 | 35 | 13.01 | 0 | 0.00 | 64 | 6.86 | 54 | 13.85 | 53 | 17.43 | 2 | 15.38 | 109 | 15.42 |
| 80+ Years | 5 | 0.78 | 4 | 1.49 | 0 | 0.00 | 9 | 0.96 | 6 | 1.54 | 11 | 3.62 | 0 | 0.00 | 17 | 2.40 |
| Not Specified | 36 | 5.58 | 2 | 0.74 | 15 | 78.95 | 53 | 5.68 | 7 | 1.79 | 3 | 0.99 | 9 | 69.23 | 19 | 2.69 |
| Total | 645 | 100.00 | 269 | 100.00 | 19 | 100.00 | **933** | 100.00 | 390 | 100.00 | 304 | 100.00 | 13 | 100.00 | **707** | 100.00 |
| **Overall (both mRNA vaccines)** |  |  |  |  |  |  |  |  |  |  |  |  |  |  |  |  |
| <6 months | 0 | 0.00 | 0 | 0.00 | 0 | 0.00 | 0 | 0.00 | 0 | 0.00 | 1 | 0.14 | 0 | 0.00 | 1 | 0.05 |
| 3-5 Years | 2 | 0.10 | 0 | 0.00 | 0 | 0.00 | 2 | 0.07 | 1 | 0.09 | 0 | 0.00 | 0 | 0.00 | 1 | 0.05 |
| 6-17 Years | 530 | 26.53 | 68 | 9.67 | 4 | 7.14 | 602 | 21.84 | 146 | 13.15 | 23 | 3.13 | 1 | 4.35 | 170 | 9.10 |
| 18-29 Years | 721 | 36.09 | 166 | 23.61 | 2 | 3.57 | 889 | 32.25 | 347 | 31.26 | 113 | 15.35 | 0 | 0.00 | 460 | 24.61 |
| 30-39 Years | 292 | 14.61 | 120 | 17.07 | 2 | 3.57 | 414 | 15.02 | 175 | 15.77 | 116 | 15.76 | 1 | 4.35 | 292 | 15.62 |
| 40-49 Years | 130 | 6.51 | 110 | 15.65 | 0 | 0.00 | 240 | 8.71 | 113 | 10.18 | 137 | 18.61 | 0 | 0.00 | 250 | 13.38 |
| 50-59 Years | 82 | 4.10 | 93 | 13.23 | 1 | 1.79 | 176 | 6.38 | 116 | 10.45 | 125 | 16.98 | 0 | 0.00 | 241 | 12.89 |
| 60-64 Years | 28 | 1.40 | 45 | 6.40 | 2 | 3.57 | 75 | 2.72 | 44 | 3.96 | 74 | 10.05 | 1 | 4.35 | 119 | 6.37 |
| 65-79 Years | 81 | 4.05 | 66 | 9.39 | 2 | 3.57 | 149 | 5.40 | 121 | 10.90 | 103 | 13.99 | 4 | 17.39 | 228 | 12.20 |
| 80+ Years | 10 | 0.50 | 8 | 1.14 | 0 | 0.00 | 18 | 0.65 | 18 | 1.62 | 20 | 2.72 | 0 | 0.00 | 38 | 2.03 |
| Not Specified | 122 | 6.11 | 27 | 3.84 | 43 | 76.79 | 192 | 6.96 | 29 | 2.61 | 24 | 3.26 | 16 | 69.57 | 69 | 3.69 |
| Total | 1998 | 100.00 | 703 | 100.00 | 56 | 100.00 | 2757 | 100.00 | 1110 | 100.00 | 736 | 100.00 | 23 | 100.00 | 1869 | 100.00 |
| **Grand Total** | **1998** | **72.47** | **703** | **25.50** | **56** | **2.03** | **2757** | **100.00** | **1110** | **59.39** | **736** | **39.38** | **23** | **1.23** | **1869** | **100.00** |

**Table 3a:** Myocarditis and pericarditis events reported to VAERS following Pfizer/BioNTech COVID-19 vaccine (Comirnaty), by dose. Datalock point 14 March 2022. Percentages per age group are presented.

| Age Group | Myocarditis |  |  |  |  |  |  |  | Pericarditis |  |  |  |  |  |  |  |
| --- | --- | --- | --- | --- | --- | --- | --- | --- | --- | --- | --- | --- | --- | --- | --- | --- |
|  | Male |  | Female |  | Sex Not Specified |  | Total |  | Male |  | Female |  | Sex Not Specified |  | Total |  |
|  | n | % | n | % | n | % | N | % | n | % | n | % | n | % | N | % |
| **Dose 1** |  |  |  |  |  |  |  |  |  |  |  |  |  |  |  |  |
| <6 months | 0 | 0.00 | 0 | 0.00 | 0 | 0.00 | 0 | 0.00 | 0 | 0.00 | 0 | 0.00 | 0 | 0.00 | 0 | 0.00 |
| 3-5 Years | 0 | 0.00 | 0 | 0.00 | 0 | 0.00 | 0 | 0.00 | 1 | 0.53 | 0 | 0.00 | 0 | 0.00 | 1 | 0.30 |
| 6-17 Years | 95 | 37.55 | 13 | 10.24 | 0 | 0.00 | 108 | 28.20 | 28 | 14.81 | 10 | 7.09 | 0 | 0.00 | 38 | 11.48 |
| 18-29 Years | 69 | 27.27 | 23 | 18.11 | 0 | 0.00 | 92 | 24.02 | 56 | 29.63 | 21 | 14.89 | 0 | 0.00 | 77 | 23.26 |
| 30-39 years | 37 | 14.62 | 28 | 22.05 | 1 | 33.33 | 66 | 17.23 | 38 | 20.11 | 26 | 18.44 | 0 | 0.00 | 64 | 19.34 |
| 40-49 years | 15 | 5.93 | 18 | 14.17 | 0 | 0.00 | 33 | 8.62 | 12 | 6.35 | 23 | 16.31 | 0 | 0.00 | 35 | 10.57 |
| 50-59 years | 8 | 3.16 | 19 | 14.96 | 0 | 0.00 | 27 | 7.05 | 21 | 11.11 | 27 | 19.15 | 0 | 0.00 | 48 | 14.50 |
| 60-64 years | 3 | 1.19 | 8 | 6.30 | 0 | 0.00 | 11 | 2.87 | 6 | 3.17 | 15 | 10.64 | 0 | 0.00 | 21 | 6.34 |
| 65-79 Years | 14 | 5.53 | 10 | 7.87 | 0 | 0.00 | 24 | 6.27 | 22 | 11.64 | 13 | 9.22 | 0 | 0.00 | 35 | 10.57 |
| 80+ Years | 0 | 0.00 | 3 | 2.36 | 0 | 0.00 | 3 | 0.78 | 3 | 1.59 | 2 | 1.42 | 0 | 0.00 | 5 | 1.51 |
| Not Specified | 12 | 4.74 | 5 | 3.94 | 2 | 66.67 | 19 | 4.96 | 2 | 1.06 | 4 | 2.84 | 1 | 100.00 | 7 | 2.11 |
| **Total** | **253** | **100.00** | **127** | **100.00** | **3** | **100.00** | **383** | **100.00** | **189** | **100.00** | **141** | **100.00** | **1** | **100.00** | **331** | **100.00** |
| **Dose 2** |  |  |  |  |  |  |  |  |  |  |  |  |  |  |  |  |
| <6 months | 0 | 0.00 | 0 | 0.00 | 0 | 0.00 | 0 | 0.00 | 0 | 0.00 | 1 | 0.59 | 0 | 0.00 | 1 | 0.19 |
| 3-5 Years | 2 | 0.27 | 0 | 0.00 | 0 | 0.00 | 2 | 0.21 | 0 | 0.00 | 0 | 0.00 | 0 | 0.00 | 0 | 0.00 |
| 6-17 Years | 305 | 40.99 | 37 | 18.05 | 1 | 14.29 | 343 | 35.88 | 83 | 23.12 | 11 | 6.51 | 0 | 0.00 | 94 | 17.67 |
| 18-29 Years | 210 | 28.23 | 47 | 22.93 | 1 | 14.29 | 258 | 26.99 | 105 | 29.25 | 15 | 8.88 | 0 | 0.00 | 120 | 22.56 |
| 30-39 years | 84 | 11.29 | 34 | 16.59 | 0 | 0.00 | 118 | 12.34 | 62 | 17.27 | 32 | 18.93 | 0 | 0.00 | 94 | 17.67 |
| 40-49 years | 41 | 5.51 | 29 | 14.15 | 0 | 0.00 | 70 | 7.32 | 27 | 7.52 | 30 | 17.75 | 0 | 0.00 | 57 | 10.71 |
| 50-59 years | 17 | 2.28 | 26 | 12.68 | 0 | 0.00 | 43 | 4.50 | 32 | 8.91 | 31 | 18.34 | 0 | 0.00 | 63 | 11.84 |
| 60-64 years | 6 | 0.81 | 6 | 2.93 | 0 | 0.00 | 12 | 1.26 | 7 | 1.95 | 17 | 10.06 | 0 | 0.00 | 24 | 4.51 |
| 65-79 Years | 21 | 2.82 | 14 | 6.83 | 0 | 0.00 | 35 | 3.66 | 28 | 7.80 | 19 | 11.24 | 1 | 25.00 | 48 | 9.02 |
| 80+ Years | 3 | 0.40 | 0 | 0.00 | 0 | 0.00 | 3 | 0.31 | 6 | 1.67 | 3 | 1.78 | 0 | 0.00 | 9 | 1.69 |
| Not Specified | 55 | 7.39 | 12 | 5.85 | 5 | 71.43 | 72 | 7.53 | 9 | 2.51 | 10 | 5.92 | 3 | 75.00 | 22 | 4.14 |
| **Total** | **744** | **100.00** | **205** | **100.00** | **7** | **100.00** | **956** | **100.00** | **359** | **100.00** | **169** | **100.00** | **4** | **100.00** | **532** | **100.00** |
| **Dose 3** |  |  |  |  |  |  |  |  |  |  |  |  |  |  |  |  |
| <6 months | 0 | 0.00 | 0 | 0.00 | 0 | 0.00 | 0 | 0.00 | 0 | 0.00 | 0 | 0.00 | 0 | 0.00 | 0 | 0.00 |
| 3-5 Years | 0 | 0.00 | 0 | 0.00 | 0 | 0.00 | 0 | 0.00 | 0 | 0.00 | 0 | 0.00 | 0 | 0.00 | 0 | 0.00 |
| 6-17 Years | 34 | 32.08 | 4 | 12.12 | 0 | 0.00 | 38 | 26.57 | 10 | 18.87 | 0 | 0.00 | 0 | 0.00 | 10 | 9.01 |
| 18-29 Years | 29 | 27.36 | 9 | 27.27 | 0 | 0.00 | 38 | 26.57 | 10 | 18.87 | 5 | 8.93 | 0 | 0.00 | 15 | 13.51 |
| 30-39 years | 13 | 12.26 | 4 | 12.12 | 0 | 0.00 | 17 | 11.89 | 5 | 9.43 | 13 | 23.21 | 0 | 0.00 | 18 | 16.22 |
| 40-49 years | 2 | 1.89 | 5 | 15.15 | 0 | 0.00 | 7 | 4.90 | 7 | 13.21 | 17 | 30.36 | 0 | 0.00 | 24 | 21.62 |
| 50-59 years | 7 | 6.60 | 4 | 12.12 | 0 | 0.00 | 11 | 7.69 | 4 | 7.55 | 7 | 12.50 | 0 | 0.00 | 11 | 9.91 |
| 60-64 years | 2 | 1.89 | 3 | 9.09 | 0 | 0.00 | 5 | 3.50 | 1 | 1.89 | 6 | 10.71 | 0 | 0.00 | 7 | 6.31 |
| 65-79 Years | 12 | 11.32 | 3 | 9.09 | 2 | 50.00 | 17 | 11.89 | 11 | 20.75 | 5 | 8.93 | 1 | 50.00 | 17 | 15.32 |
| 80+ Years | 2 | 1.89 | 0 | 0.00 | 0 | 0.00 | 2 | 1.40 | 3 | 5.66 | 1 | 1.79 | 0 | 0.00 | 4 | 3.60 |
| Not Specified | 5 | 4.72 | 1 | 3.03 | 2 | 50.00 | 8 | 5.59 | 2 | 3.77 | 2 | 3.57 | 1 | 50.00 | 5 | 4.50 |
| **Total** | **106** | **100.00** | **33** | **100.00** | **4** | **100.00** | **143** | **100.00** | **53** | **100.00** | **56** | **100.00** | **2** | **100.00** | **111** | **100.00** |
| **Dose 4+** |  |  |  |  |  |  |  |  |  |  |  |  |  |  |  |  |
| 60-64 Years | 0 | 0.00 | 0 | 0.00 | 0 | 0.00 | 0 | 0.00 | 0 | 0.00 | 2 | 100.00 | 0 | 0.00 | 2 | 100.00 |
| **Total** | **0** | **0.00** | **0** | **0.00** | **0** | **0.00** | **0** | **0.00** | **0** | **0.00** | **2** | **100.00** | **0** | **0.00** | **2** | **100.00** |
| **Unknown dose** |  |  |  |  |  |  |  |  |  |  |  |  |  |  |  |  |
| <6 months | 0 | 0.00 | 0 | 0.00 | 0 | 0.00 | 0 | 0.00 | 0 | 0.00 | 0 | 0.00 | 0 | 0.00 | 0 | 0.00 |
| 3-5 Years | 0 | 0.00 | 0 | 0.00 | 0 | 0.00 | 0 | 0.00 | 0 | 0.00 | 0 | 0.00 | 0 | 0.00 | 0 | 0.00 |
| 6-17 Years | 91 | 36.40 | 13 | 18.84 | 1 | 4.35 | 105 | 30.70 | 24 | 20.17 | 2 | 3.13 | 1 | 25.00 | 27 | 14.44 |
| 18-29 Years | 96 | 38.40 | 23 | 33.33 | 0 | 0.00 | 119 | 34.80 | 46 | 38.66 | 10 | 15.63 | 0 | 0.00 | 56 | 29.95 |
| 30-39 years | 23 | 9.20 | 3 | 4.35 | 1 | 4.35 | 27 | 7.89 | 11 | 9.24 | 7 | 10.94 | 0 | 0.00 | 18 | 9.63 |
| 40-49 years | 10 | 4.00 | 7 | 10.14 | 0 | 0.00 | 17 | 4.97 | 10 | 8.40 | 13 | 20.31 | 0 | 0.00 | 23 | 12.30 |
| 50-59 years | 9 | 3.60 | 7 | 10.14 | 0 | 0.00 | 16 | 4.68 | 7 | 5.88 | 8 | 12.50 | 0 | 0.00 | 15 | 8.02 |
| 60-64 years | 2 | 0.80 | 4 | 5.80 | 2 | 8.70 | 8 | 2.34 | 6 | 5.04 | 3 | 4.69 | 0 | 0.00 | 9 | 4.81 |
| 65-79 Years | 5 | 2.00 | 4 | 5.80 | 0 | 0.00 | 9 | 2.63 | 6 | 5.04 | 13 | 20.31 | 1 | 25.00 | 20 | 10.70 |
| 80+ Years | 0 | 0.00 | 1 | 1.45 | 0 | 0.00 | 1 | 0.29 | 0 | 0.00 | 3 | 4.69 | 0 | 0.00 | 3 | 1.60 |
| Not Specified | 14 | 5.60 | 7 | 10.14 | 19 | 82.61 | 40 | 11.70 | 9 | 7.56 | 5 | 7.81 | 2 | 50.00 | 16 | 8.56 |
| Total | 250 | 100.00 | 69 | 100.00 | 23 | 100.00 | 342 | 100.00 | 119 | 100.00 | 64 | 100.00 | 4 | 100.00 | 187 | 100.00 |
| Grand Total | 1353 |  | 434 |  | 37 |  | **1824** |  | 720 |  | 432 |  | 11 |  | **1162** |  |

**Table 3b:** Myocarditis and pericarditis events reported to VAERS following Moderna COVID-19 vaccine (Spikevax), by dose. Datalock point 14 March 2022. Percentages per age group are presented.

| Age Group | Myocarditis |  |  |  |  |  |  |  | Pericarditis |  |  |  |  |  |  |  |
| --- | --- | --- | --- | --- | --- | --- | --- | --- | --- | --- | --- | --- | --- | --- | --- | --- |
|  | Male |  | Female |  | Sex Not Specified |  | Total |  | Male |  | Female |  | Sex Not Specified |  | Total |  |
|  | n | % | n | % | n | % | N | % | n | % | n | % | n | % | N | % |
| **Dose 1** |  |  |  |  |  |  |  |  |  |  |  |  |  |  |  |  |
| <6 months | 0 | 0.00 | 0 | 0.00 | 0 | 0.00 | 0 | 0.00 | 0 | 0.00 | 0 | 0.00 | 0 | 0.00 | 0 | 0.00 |
| 3-5 Years | 0 | 0.00 | 0 | 0.00 | 0 | 0.00 | 0 | 0.00 | 0 | 0.00 | 0 | 0.00 | 0 | 0.00 | 0 | 0.00 |
| 6-17 Years | 1 | 0.56 | 0 | 0.00 | 0 | 0.00 | 1 | 0.35 | 0 | 0.00 | 0 | 0.00 | 0 | 0.00 | 0 | 0.00 |
| 18-29 Years | 74 | 41.11 | 18 | 18.00 | 0 | 0.00 | 92 | 32.17 | 33 | 32.35 | 23 | 20.00 | 0 | 0.00 | 56 | 25.23 |
| 30-39 years | 42 | 23.33 | 18 | 18.00 | 0 | 0.00 | 60 | 20.98 | 15 | 14.71 | 17 | 14.78 | 0 | 0.00 | 32 | 14.41 |
| 40-49 years | 16 | 8.89 | 21 | 21.00 | 0 | 0.00 | 37 | 12.94 | 21 | 20.59 | 20 | 17.39 | 0 | 0.00 | 41 | 18.47 |
| 50-59 years | 15 | 8.33 | 16 | 16.00 | 1 | 16.67 | 32 | 11.19 | 12 | 11.76 | 19 | 16.52 | 0 | 0.00 | 31 | 13.96 |
| 60-64 years | 3 | 1.67 | 15 | 15.00 | 0 | 0.00 | 18 | 6.29 | 7 | 6.86 | 12 | 10.43 | 0 | 0.00 | 19 | 8.56 |
| 65-79 Years | 10 | 5.56 | 11 | 11.00 | 0 | 0.00 | 21 | 7.34 | 11 | 10.78 | 16 | 13.91 | 2 | 40.00 | 29 | 13.06 |
| 80+ Years | 2 | 1.11 | 1 | 1.00 | 0 | 0.00 | 3 | 1.05 | 0 | 0.00 | 5 | 4.35 | 0 | 0.00 | 5 | 2.25 |
| Not Specified | 17 | 9.44 | 0 | 0.00 | 5 | 83.33 | 22 | 7.69 | 3 | 2.94 | 3 | 2.61 | 3 | 60.00 | 9 | 4.05 |
| Total | 180 | 100.00 | 100 | 100.00 | 6 | 100.00 | 286 | 100.00 | 102 | 100.00 | 115 | 100.00 | 5 | 100.00 | 222 | 100.00 |
| **Dose 2** |  |  |  |  |  |  |  |  |  |  |  |  |  |  |  |  |
| <6 months | 0 | 0.00 | 0 | 0.00 | 0 | 0.00 | 0 | 0.00 | 0 | 0.00 | 0 | 0.00 | 0 | 0.00 | 0 | 0.00 |
| 3-5 Years | 0 | 0.00 | 0 | 0.00 | 0 | 0.00 | 0 | 0.00 | 0 | 0.00 | 0 | 0.00 | 0 | 0.00 | 0 | 0.00 |
| 6-17 Years | 2 | 0.72 | 0 | 0.00 | 0 | 0.00 | 2 | 0.52 | 1 | 0.55 | 0 | 0.00 | 0 | 0.00 | 1 | 0.33 |
| 18-29 Years | 152 | 54.48 | 29 | 29.00 | 1 | 20.00 | 182 | 47.40 | 63 | 34.81 | 24 | 19.67 | 0 | 0.00 | 87 | 28.52 |
| 30-39 years | 57 | 20.43 | 15 | 15.00 | 0 | 0.00 | 72 | 18.75 | 30 | 16.57 | 11 | 9.02 | 0 | 0.00 | 41 | 13.44 |
| 40-49 years | 24 | 8.60 | 24 | 24.00 | 0 | 0.00 | 48 | 12.50 | 20 | 11.05 | 21 | 17.21 | 0 | 0.00 | 41 | 13.44 |
| 50-59 years | 20 | 7.17 | 13 | 13.00 | 0 | 0.00 | 33 | 8.59 | 25 | 13.81 | 24 | 19.67 | 0 | 0.00 | 49 | 16.07 |
| 60-64 years | 6 | 2.15 | 5 | 5.00 | 0 | 0.00 | 11 | 2.86 | 10 | 5.52 | 12 | 9.84 | 0 | 0.00 | 22 | 7.21 |
| 65-79 Years | 13 | 4.66 | 12 | 12.00 | 0 | 0.00 | 25 | 6.51 | 26 | 14.36 | 26 | 21.31 | 0 | 0.00 | 52 | 17.05 |
| 80+ Years | 1 | 0.36 | 1 | 1.00 | 0 | 0.00 | 2 | 0.52 | 4 | 2.21 | 4 | 3.28 | 0 | 0.00 | 8 | 2.62 |
| Not Specified | 4 | 1.43 | 1 | 1.00 | 4 | 80.00 | 9 | 2.34 | 2 | 1.10 | 0 | 0.00 | 2 | 100.00 | 4 | 1.31 |
| Total | 279 | 100.00 | 100 | 100.00 | 5 | 100.00 | 384 | 100.00 | 181 | 100.00 | 122 | 100.00 | 2 | 100.00 | 305 | 100.00 |
| **Dose 3** |  |  |  |  |  |  |  |  |  |  |  |  |  |  |  |  |
| <6 months | 0 | 0.00 | 0 | 0.00 | 0 | 0.00 | 0 | 0.00 | 0 | 0.00 | 0 | 0.00 | 0 | 0.00 | 0 | 0.00 |
| 3-5 Years | 0 | 0.00 | 0 | 0.00 | 0 | 0.00 | 0 | 0.00 | 0 | 0.00 | 0 | 0.00 | 0 | 0.00 | 0 | 0.00 |
| 6-17 Years | 1 | 1.96 | 0 | 0.00 | 0 | 0.00 | 1 | 1.16 | 0 | 0.00 | 0 | 0.00 | 0 | 0.00 | 0 | 0.00 |
| 18-29 Years | 29 | 56.86 | 8 | 22.86 | 0 | 0.00 | 37 | 43.02 | 10 | 29.41 | 6 | 16.22 | 0 | 0.00 | 16 | 22.54 |
| 30-39 years | 9 | 17.65 | 10 | 28.57 | 0 | 0.00 | 19 | 22.09 | 4 | 11.76 | 7 | 18.92 | 0 | 0.00 | 11 | 15.49 |
| 40-49 years | 6 | 11.76 | 4 | 11.43 | 0 | 0.00 | 10 | 11.63 | 5 | 14.71 | 8 | 21.62 | 0 | 0.00 | 13 | 18.31 |
| 50-59 years | 2 | 3.92 | 4 | 11.43 | 0 | 0.00 | 6 | 6.98 | 7 | 20.59 | 5 | 13.51 | 0 | 0.00 | 12 | 16.90 |
| 60-64 years | 1 | 1.96 | 2 | 5.71 | 0 | 0.00 | 3 | 3.49 | 1 | 2.94 | 3 | 8.11 | 0 | 0.00 | 4 | 5.63 |
| 65-79 Years | 3 | 5.88 | 6 | 17.14 | 0 | 0.00 | 9 | 10.47 | 6 | 17.65 | 6 | 16.22 | 0 | 0.00 | 12 | 16.90 |
| 80+ Years | 0 | 0.00 | 1 | 2.86 | 0 | 0.00 | 1 | 1.16 | 1 | 2.94 | 2 | 5.41 | 0 | 0.00 | 3 | 4.23 |
| Not Specified | 0 | 0.00 | 0 | 0.00 | 0 | 0.00 | 0 | 0.00 | 0 | 0.00 | 0 | 0.00 | 0 | 0.00 | 0 | 0.00 |
| Total | 51 | 100.00 | 35 | 100.00 | 0 | 0.00 | 86 | 100.00 | 34 | 100.00 | 37 | 100.00 | 0 | 0.00 | 71 | 100.00 |
| **Dose 4+** |  |  |  |  |  |  |  |  |  |  |  |  |  |  |  |  |
| 40-49 Years | 0 | 0.00 | 1 | 100.00 | 0 | 0.00 | 1 | 100.00 | 0 | 0.00 | 0 | 0.00 | 0 | 0.00 | 0 | 0.00 |
| Total | 0 | 0.00 | 1 | 100.00 | 0 | 0.00 | 1 | 100.00 | 0 | 0.00 | 0 | 0.00 | 0 | 0.00 | 0 | 0.00 |
| **Unknown dose** |  |  |  |  |  |  |  |  |  |  |  |  |  |  |  |  |
| <6 months | 0 | 0.00 | 0 | 0.00 | 0 | 0.00 | 0 | 0.00 | 0 | 0.00 | 0 | 0.00 | 0 | 0.00 | 0 | 0.00 |
| 3-5 Years | 0 | 0.00 | 0 | 0.00 | 0 | 0.00 | 0 | 0.00 | 0 | 0.00 | 0 | 0.00 | 0 | 0.00 | 0 | 0.00 |
| 6-17 Years | 1 | 0.74 | 1 | 3.03 | 2 | 25.00 | 4 | 2.27 | 0 | 0.00 | 0 | 0.00 | 0 | 0.00 | 0 | 0.00 |
| 18-29 Years | 62 | 45.93 | 9 | 27.27 | 0 | 0.00 | 71 | 40.34 | 24 | 32.88 | 9 | 30.00 | 0 | 0.00 | 33 | 30.28 |
| 30-39 years | 27 | 20.00 | 8 | 24.24 | 0 | 0.00 | 35 | 19.89 | 10 | 13.70 | 3 | 10.00 | 1 | 16.67 | 14 | 12.84 |
| 40-49 years | 16 | 11.85 | 1 | 3.03 | 0 | 0.00 | 17 | 9.66 | 11 | 15.07 | 5 | 16.67 | 0 | 0.00 | 16 | 14.68 |
| 50-59 years | 4 | 2.96 | 4 | 12.12 | 0 | 0.00 | 8 | 4.55 | 8 | 10.96 | 4 | 13.33 | 0 | 0.00 | 12 | 11.01 |
| 60-64 years | 5 | 3.70 | 2 | 6.06 | 0 | 0.00 | 7 | 3.98 | 6 | 8.22 | 4 | 13.33 | 1 | 16.67 | 11 | 10.09 |
| 65-79 Years | 3 | 2.22 | 6 | 18.18 | 0 | 0.00 | 9 | 5.11 | 11 | 15.07 | 5 | 16.67 | 0 | 0.00 | 16 | 14.68 |
| 80+ Years | 2 | 1.48 | 1 | 3.03 | 0 | 0.00 | 3 | 1.70 | 1 | 1.37 | 0 | 0.00 | 0 | 0.00 | 1 | 0.92 |
| Not Specified | 15 | 11.11 | 1 | 3.03 | 6 | 75.00 | 22 | 12.50 | 2 | 2.74 | 0 | 0.00 | 4 | 66.67 | 6 | 5.50 |
| Total | 135 | 100.00 | 33 | 100.00 | 8 | 100.00 | 176 | 100.00 | 73 | 100.00 | 30 | 100.00 | 6 | 100.00 | 109 | 100.00 |
| Grand Total | 645 |  | 269 |  | 19 |  | **933** |  | 390 |  | 304 |  | 13 |  | **707** |  |

The EudraVigilance database contained the highest total reports of events with 9211 events reported following Comirnaty and 2786 following Spikevax (Table 4; Figure 2A). There had been approximately 296.05 million vaccinees who had received at least one dose of Comirnaty in the EU/EEA. Therefore, the reporting rates were calculated as 14.50 reports of myocarditis and 16.61 reports of pericarditis per million Comirnaty recipients, giving a combined reporting rate of 15.56 cases of myocarditis and pericarditis per million people who received at least one dose of Comirnaty (Figure 2B). For Spikevax, there had been approximately 46.56 million first doses of Spikevax administered in the EU and EEA. Thus, reporting rates in the EU/EEA are currently 28.01 reports of myocarditis and 31.83 reports of pericarditis per million vaccinees, giving a combined reporting rate of 29.92 per million Spikevax recipients (Figure 2B).

**Table 4:** Myocarditis and pericarditis events reported to EudraVigilance for the European Economic Area. Datalock point 14 March 2022. Percentages per age group are presented.

| Age Group | Myocarditis |  |  |  |  |  |  |  | Pericarditis |  |  |  |  |  |  |  |
| --- | --- | --- | --- | --- | --- | --- | --- | --- | --- | --- | --- | --- | --- | --- | --- | --- |
|  | Male |  | Female |  | Sex Not Specified |  | Total |  | Male |  | Female |  | Sex Not Specified |  | Total |  |
|  | n | % | n | % | n | % | N | % | n | % | n | % | n | % | N | % |
| **Comirnaty (Pfizer/BioNTech)** |  |  |  |  |  |  |  |  |  |  |  |  |  |  |  |  |
| 2 Months-2 Years | 0 | 0.00 | 0 | 0.00 | 0 | 0.00 | 0 | 0.00 | 67 | 63.81 | 38 | 36.19 | 0 | 0.00 | 105 | 100.00 |
| 3-11 Years | 4 | 66.67 | 2 | 33.33 | 0 | 0.00 | 6 | 100.00 | 798 | 48.33 | 848 | 51.36 | 5 | 0.30 | 1651 | 100.00 |
| 12-17 Years | 507 | 86.82 | 72 | 12.33 | 5 | 0.86 | 584 | 100.00 | 164 | 56.75 | 125 | 43.25 | 0 | 0.00 | 289 | 100.00 |
| 18-64 Years | 2302 | 70.16 | 956 | 29.14 | 23 | 0.70 | 3281 | 100.00 | 307 | 46.30 | 351 | 52.94 | 5 | 0.75 | 663 | 100.00 |
| 65-85 Years | 130 | 50.58 | 125 | 48.64 | 2 | 0.78 | 257 | 100.00 | 54 | 46.55 | 62 | 53.45 | 0 | 0.00 | 116 | 100.00 |
| 85+ Years | 12 | 52.17 | 11 | 47.83 | 0 | 0.00 | 23 | 100.00 | 1042 | 50.27 | 1026 | 49.49 | 5 | 0.24 | 2073 | 100.00 |
| Not Specified | 86 | 60.56 | 39 | 27.46 | 17 | 11.97 | 142 | 100.00 | 8 | 38.10 | 11 | 52.38 | 2 | 9.52 | 21 | 100.00 |
| Total | 3041 | 70.84 | 1205 | 28.07 | 47 | 1.09 | **4293** | **100.00** | 2440 | 49.61 | 2461 | 50.04 | 17 | 0.35 | **4918** | **100.00** |
| **Spikevax (Moderna)** |  |  |  |  |  |  |  |  |  |  |  |  |  |  |  |  |
| 2 Months-2 Years | 0 | 0.00 | 0 | 0.00 | 0 | 0.00 | 0 | 0.00 | 8 | 0.00 | 4 | 0.00 | 0 | 0.00 | 12 | 100.00 |
| 3-11 Years | 0 | 0.00 | 0 | 0.00 | 0 | 0.00 | 0 | 0.00 | 343 | 0.00 | 287 | 0.00 | 5 | 0.79 | 635 | 100.00 |
| 12-17 Years | 69 | 92.00 | 6 | 8.00 | 0 | 0.00 | 75 | 100.00 | 39 | 0.00 | 45 | 0.00 | 0 | 0.00 | 84 | 100.00 |
| 18-64 Years | 913 | 79.39 | 231 | 20.09 | 6 | 0.52 | 1150 | 100.00 | 0 | 0.00 | 2 | 100.00 | 0 | 0.00 | 2 | 100.00 |
| 65-85 Years | 27 | 48.21 | 29 | 51.79 | 0 | 0.00 | 56 | 100.00 | 4 | 50.00 | 4 | 50.00 | 0 | 0.00 | 8 | 100.00 |
| 85+ Years | 1 | 33.33 | 2 | 66.67 | 0 | 0.00 | 3 | 100.00 | 394 | 53.17 | 342 | 46.15 | 5 | 0.67 | 741 | 100.00 |
| Not Specified | 11 | 55.00 | 7 | 35.00 | 2 | 10.00 | 20 | 100.00 | 0 | 0.00 | 0 | 0.00 | 0 | 0.00 | 0 | - |
| **Total** | **1021** | **78.30** | **275** | **21.09** | **8** | **0.61** | **1304** | **100.00** | **788** | **53.17** | **684** | **46.15** | **10** | **0.67** | **1482** | **100.00** |
| **Overall (both mRNA vaccines)** |  |  |  |  |  |  |  |  |  |  |  |  |  |  |  |  |
| 2 Months-2 Years | 0 | 0.00 | 0 | 0.00 | 0 | 0.00 | 0 | - | 75 | 64.10 | 42 | 35.90 | 0 | 0.00 | 117 | 100.00 |
| 3-11 Years | 4 | 66.67 | 2 | 33.33 | 0 | 0.00 | 6 | 100.00 | 1141 | 49.91 | 1135 | 49.65 | 10 | 0.44 | 2286 | 100.00 |
| 12-17 Years | 576 | 87.41 | 78 | 11.84 | 5 | 0.76 | 659 | 100.00 | 203 | 54.42 | 170 | 45.58 | 0 | 0.00 | 373 | 100.00 |
| 18-64 Years | 3215 | 72.56 | 1187 | 26.79 | 29 | 0.65 | 4431 | 100.00 | 307 | 46.17 | 353 | 53.08 | 5 | 0.75 | 665 | 100.00 |
| 65-85 Years | 157 | 50.16 | 154 | 49.20 | 2 | 0.64 | 313 | 100.00 | 58 | 46.77 | 66 | 53.23 | 0 | 0.00 | 124 | 100.00 |
| 85+ Years | 13 | 50.00 | 13 | 50.00 | 0 | 0.00 | 26 | 100.00 | 1436 | 51.03 | 1368 | 48.61 | 10 | 0.36 | 2814 | 100.00 |
| Not Specified | 97 | 59.88 | 46 | 28.40 | 19 | 11.73 | 162 | 100.00 | 8 | 38.10 | 11 | 52.38 | 2 | 9.52 | 21 | 100.00 |
| **Total** | **4058** | **72.55** | **1480** | **26.46** | **55** | **0.98** | **5593** | **100.00** | **3228** | **50.44** | **3145** | **49.14** | **27** | **0.42** | **6400** | **100.00** |

In total, there were 13,573 events of myocarditis and/or pericarditis reported in the observational studies identified by systematic review of the literature.

While reporting rates for myocarditis and pericarditis have differed between the spontaneous reporting databases, overall, they demonstrate that these events are very rare (defined as occurring at a rate of <1 in 10,000) (23).

### 3.3 Fatalities following myocarditis and pericarditis after COVID-19 mRNA vaccines

There have been cases of myocarditis and pericarditis with a fatal outcome reported to spontaneous reporting systems and in the literature. There were four fatalities in the UK (0.25% of all spontaneous reports), 62 in the US (1.3% of all reports) and 56 in the EU/EEA (0.6% of all reports). Where age of fatal cases was reported (EudraVigilance and VAERS databases only), 85.83% (n=103) of fatal cases overall were aged 18 years or older. Five cases (4.17%) were aged under 18 years; all were myocarditis events (6.41% of all fatal myocarditis events reported). Ten percent of fatal cases had age unspecified. All fatal cases of pericarditis reported to EudraVigilance and VAERS were aged 18 years or older.

Fatal cases were reported in five of the 32 included studies identified by systematic literature review (Table 6) (24-28). Overall, 0.22% (n=30) of 13,571 myocarditis or pericarditis events reported in the literature had a fatal outcome (range 0.41-45.85%; Table 6). Characteristics of fatal cases were specified in one study (24). In this study, results indicated that fatal cases of myocarditis and pericarditis occurred in the adult population. There were 15 fatal myocarditis events (median age 60 [Interquartile Range (IQR) 56-78] years), 5 fatal pericarditis events (median age 71 [IQR 67-77] years), and two fatal myopericarditis events (aged 55 and 83 years).

**Table 5:**
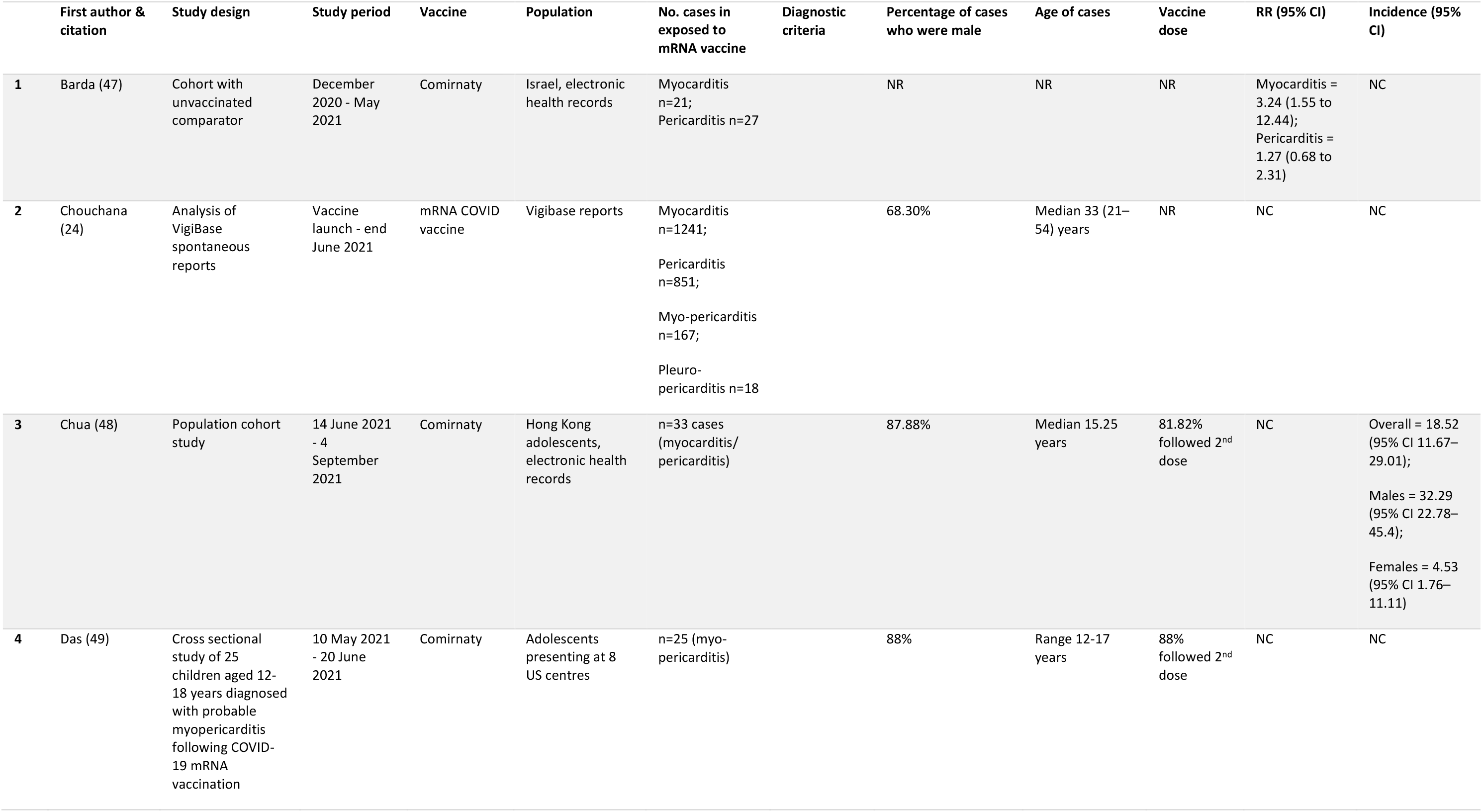
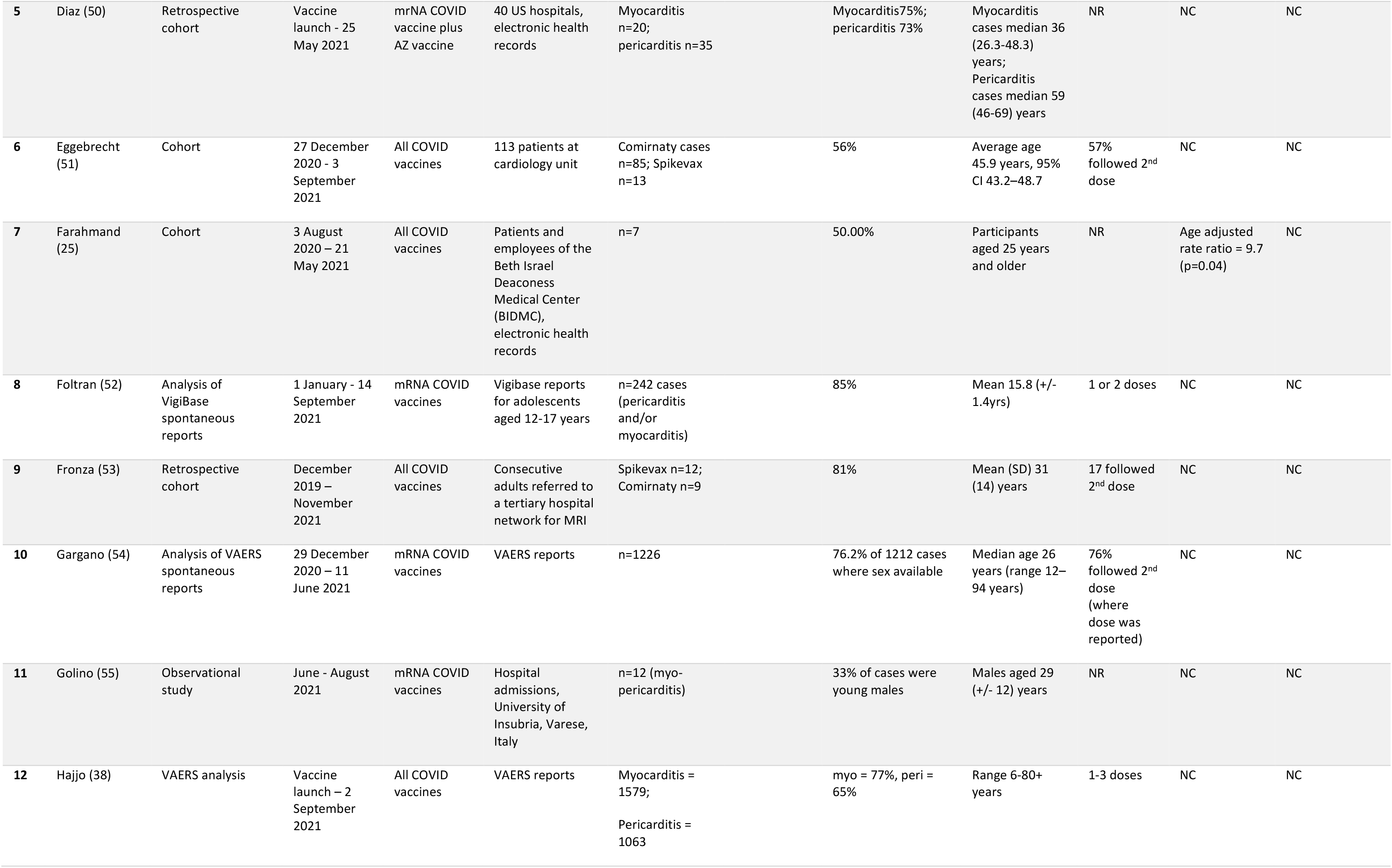
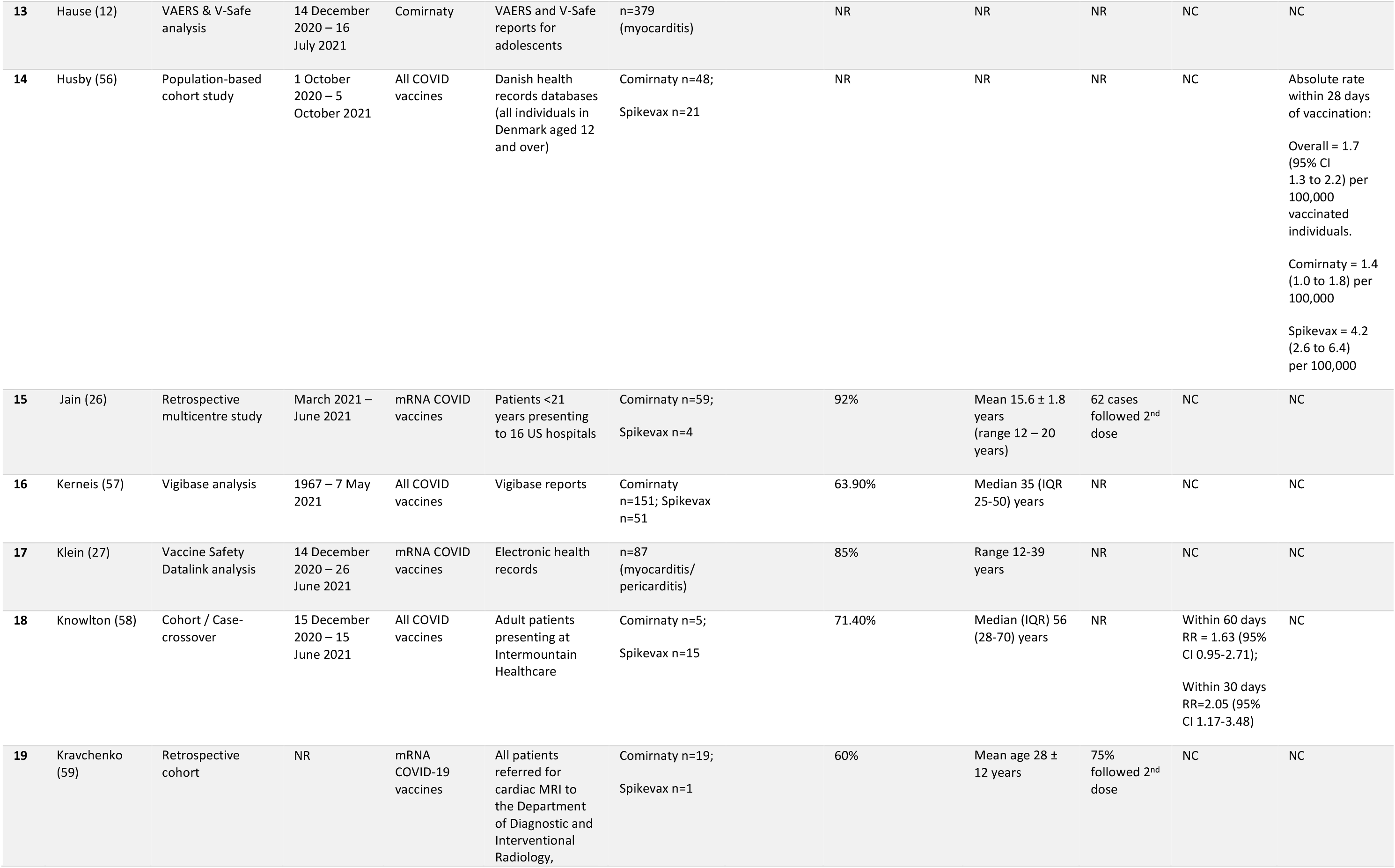
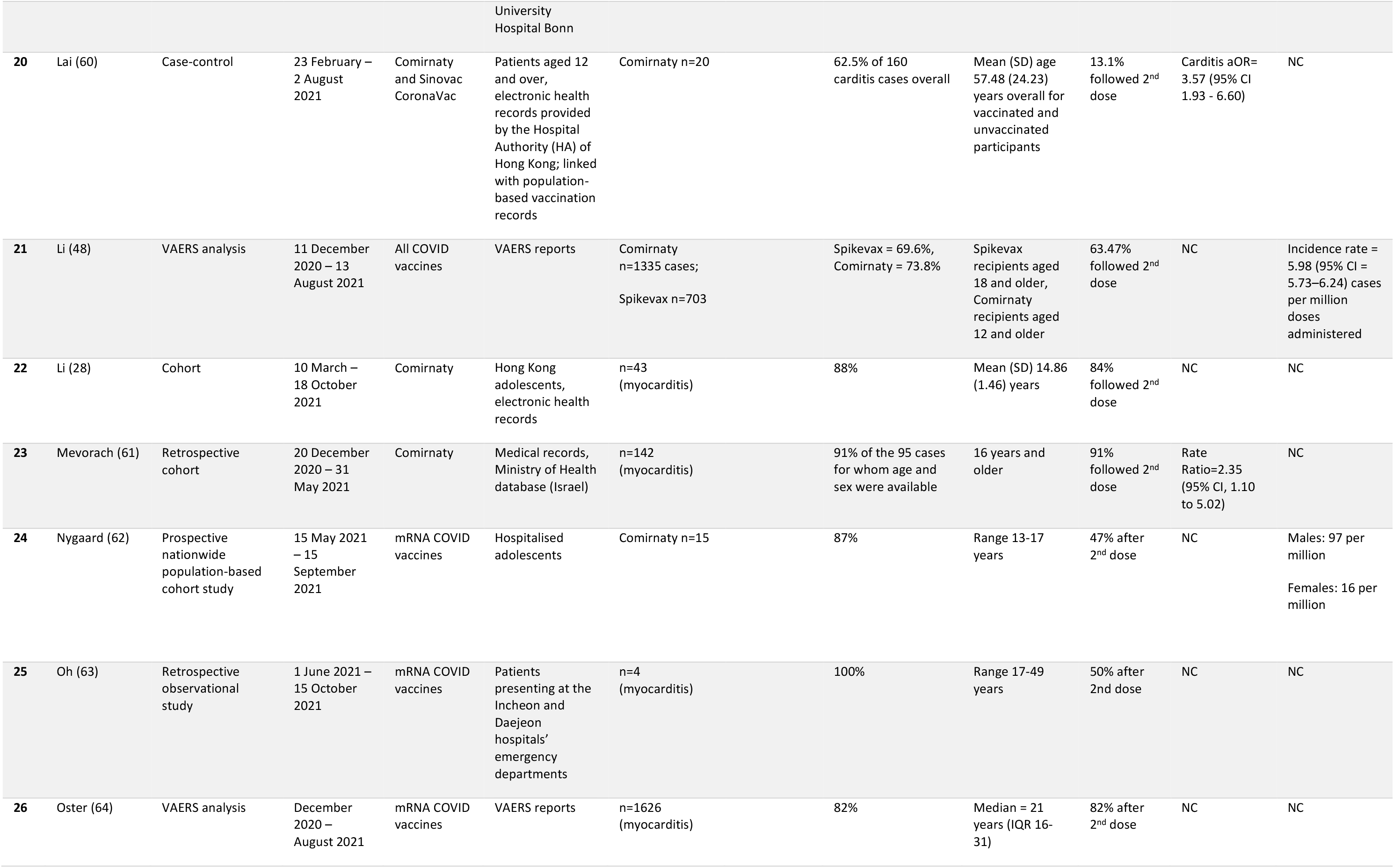
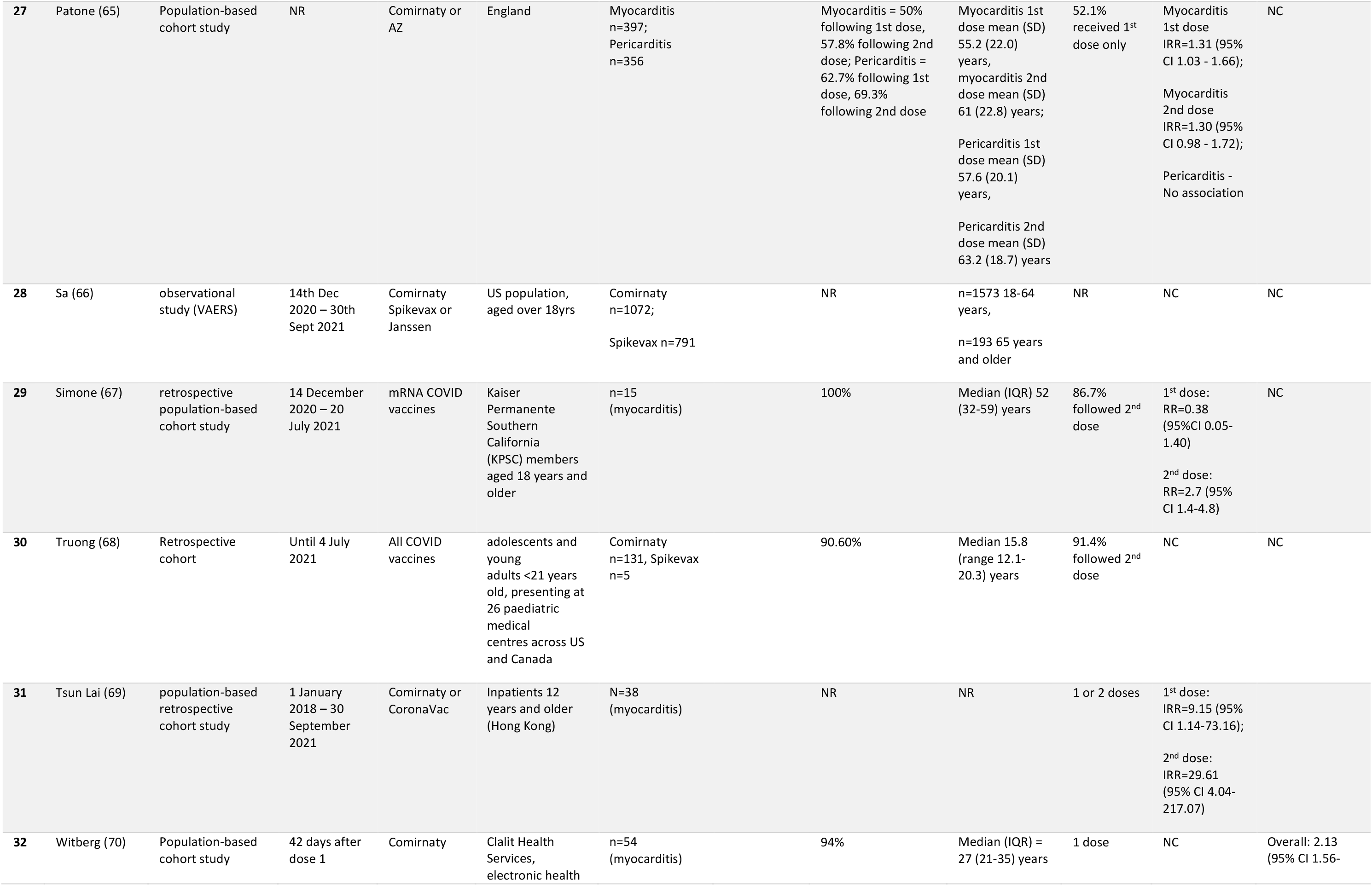
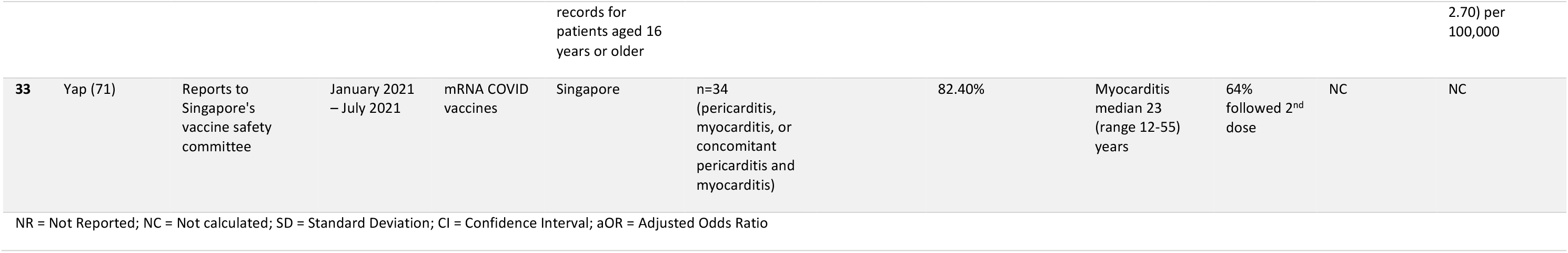
Details of each study identified by systematic review which met the inclusion criteria.

**Table 6:** Results of studies identified by systematic review, including the number of events, fatal cases, sex, and vaccine dose.

|  | Myocarditis |  | Myopericarditis |  | Pericarditis |  | Total cases |  | Fatal cases |  | Males (where specified) |  | Females (where specified) |  | Single dose |  | Two doses |  | Dose unspecified |  |
| --- | --- | --- | --- | --- | --- | --- | --- | --- | --- | --- | --- | --- | --- | --- | --- | --- | --- | --- | --- | --- |
|  | n | % | n | % | n | % | n | % | n | % | n | % | n | % | n | % | n | % | n | % |
| **1** | 21 | 43.75 | 27 | 56.25 | 0 | 0.00 | 48 | 100.00 | NR |  | NR |  | NR |  | NR |  | NR |  | NR |  |
| **2** | 1241 | 54.94 | 851 | 37.67 | 167 | 7.39 | 2259 | 100.00 | 22 | 45.83 | 1555 | 68.84 | 722 | 31.96 | NR |  | NR |  | NR |  |
| **3** | 33 | 100.00 | 0 | 0.00 | 0 | 0.00 | 33 | 100.00 | NR |  | 29 | 87.88 | 4 | 12.12 | 6 | 18.18 | 27 | 81.82 | 0 | 0 |
| **4** | 20 | 36.36 | 0 | 0.00 | 35 | 63.64 | 55 | 100.00 | NR |  | 42 | 76.36 | 15 | 27.27 | NR |  | NR |  | NR |  |
| **5** | 0 | 0.00 | 4 | 100.00 | 0 | 0.00 | 4 | 100.00 | NR |  | 2 | 50 | 2 | 50 | 3 | 75 | 1 | 25 | 0 | 0 |
| **6** | 0 | 0.00 | 10 | 100.00 | 0 | 0.00 | 10 | 100.00 | NR |  | 6 | 60 | 6 | 60 | 3 | 30 | 7 | 70 | 0 | 0 |
| **7** | 191 | 78.93 | 2 | 0.83 | 49 | 20.25 | 242 | 100.00 | 1 | 0.41 | 205 | 84.71 | 37 | 15.29 | 37 | 15.29 | 68 | 28.1 | 139 | 57.44 |
| **8** | 21 | 100.00 | 0 | 0.00 | 0 | 0.00 | 21 | 100.00 | NR |  | 17 | 80.95 | 4 | 19.05 | NR |  | NR |  | NR |  |
| **9** | 1226 | 100.00 | 0 | 0.00 | 0 | 0.00 | 1226 | 100.00 | 0 | 0.00 | 923 | 75.29 | 289 | 23.57 | 263 | 21.45 | 831 | 67.78 | 0 | 0 |
| **10** | 0 | 0.00 | 12 | 100.00 | 0 | 0.00 | 12 | 100.00 | 0 | 0.00 | NR |  | NR |  | NR |  | NR |  | NR |  |
| **11** | 1579 | 59.77 | 0 | 0.00 | 1063 | 40.23 | 2642 | 100.00 | NR |  | 1906 | 72.14 | 736 | 27.86 | NR |  | NR |  | NR |  |
| **12** | 397 | 100.00 | 0 | 0.00 | 0 | 0.00 | 397 | 100.00 | 0 | 0.00 | NR |  | NR |  | NR |  | NR |  | NR |  |
| **13** | 48 | 69.57 | 21 | 30.43 | 0 | 0.00 | 69 | 100.00 | NR |  | NR |  | NR |  | NR |  | NR |  | NR |  |
| **14** | 63 | 100.00 | 0 | 0.00 | 0 | 0.00 | 63 | 100.00 | NR |  | 58 | 92.06 | 5 | 7.94 | 1 | 1.59 | 62 | 98.41 | 0 | 0 |
| **15** | 156 | 100.00 | 0 | 0.00 | 0 | 0.00 | 156 | 100.00 | 5 | 3.21 | 131 | 83.97 | 74 | 47.44 | NR |  | NR |  | NR |  |
| **16** | 0 | 0.00 | 34 | 100.00 | 0 | 0.00 | 34 | 100.00 | NR |  | 29 | 85.29 | 5 | 14.71 | 9 | 26.47 | 24 | 70.59 | 1 | 2.94 |
| **17** | 0 | 0.00 | 20 | 100.00 | 0 | 0.00 | 20 | 100.00 | 1 | 5.00 | 15 | 75 | 6 | 30 | 13 | 65 | 7 | 35 | 0 | 0 |
| **18** | 20 | 100.00 | 0 | 0.00 | 0 | 0.00 | 20 | 100.00 | NR |  | 12 | 60 | 8 | 40 | 5 | 25 | 15 | 75 | 0 | 0 |
| **19** | 0 | 0.00 | 20 | 100.00 | 0 | 0.00 | 20 | 100.00 | NR |  | NR |  | NR |  | NR |  | NR |  | NR |  |
| **20** | 0 | 0.00 | 2038 | 100.00 | 0 | 0.00 | 2038 | 100.00 | NR |  | 1474 | 72.33 | 535 | 26.25 | NR |  | NR |  | NR |  |
| **21** | 43 | 100.00 | 0 | 0.00 | 0 | 0.00 | 43 | 100.00 | NR |  | 38 | 88.37 | 5 | 11.63 | 7 | 16.28 | 36 | 83.72 | 0 | 0 |
| **22** | 182 | 100.00 | 0 | 0.00 | 0 | 0.00 | 182 | 100.00 | 1 | 0.55 | NR |  | NR |  | 0 | 0 | 142 | 78.02 | 40 | 21.98 |
| **23** | 0 | 0.00 | 12 | 80.00 | 3 | 20.00 | 15 | 100.00 | 0 | 0.00 | 13 | 86.67 | 2 | 13.33 | 8 | 53.33 | 7 | 46.67 | 0 | 0 |
| **24** | 4 | 100.00 | 0 | 0.00 | 0 | 0.00 | 4 | 100.00 | 0 | 0.00 | 4 | 100 | 0 | 0 | 2 | 50 | 2 | 50 | 0 | 0 |
| **25** | 1625 | 70.38 | 0 | 0.00 | 684 | 29.62 | 2309 | 100.00 | 0 | 0.00 | 1334 | 57.77 | 290 | 12.56 | 273 | 11.82 | 1265 | 54.79 | 0 | 0 |
| **26** | 158 | 54.11 | 0 | 0.00 | 134 | 45.89 | 292 | 100.00 | NR |  | 173 | 59.25 | 119 | 40.75 | 153 | 52.4 | 139 | 47.6 | 0 | 0 |
| **27** | 0 | 0.00 | 1072 | 100.00 | 0 | 0.00 | 1072 | 100.00 | NR |  | NR |  | NR |  | NR |  | NR |  | NR |  |
| **28** | 15 | 100.00 | 0 | 0.00 | 0 | 0.00 | 15 | 100.00 | NR |  | 15 | 100 | 0 | 0 | 2 | 13.33 | 13 | 86.67 | 0 | 0 |
| **29** | 146 | 100.00 | 0 | 0.00 | 0 | 0.00 | 146 | 100.00 | NR |  | 126 | 86.3 | 20 | 13.7 | 12 | 8.22 | 134 | 91.78 | 0 | 0 |
| **30** | 38 | 100.00 | 0 | 0.00 | 0 | 0.00 | 38 | 100.00 | NR |  | NR |  | NR |  | 8 | 21.05 | 30 | 78.95 | 0 | 0 |
| **31** | 54 | 100.00 | 0 | 0.00 | 0 | 0.00 | 54 | 100.00 | NR |  | 51 | 94.44 | 3 | 5.56 | 17 | 31.48 | 37 | 68.52 | 0 | 0 |
| **32** | 14 | 41.18 | 11 | 32.35 | 9 | 26.47 | 34 | 100.00 | 0 | 0.00 | 28 | 82.35 | 6 | 17.65 | 15 | 44.12 | 19 | 55.88 | 0 | 0 |
| **Total** | 7295 |  | 4134 |  | 2144 |  | 13573 |  | 30 |  | 8186 |  | 2893 |  | 837 |  | 2866 |  | 180 |  |
| **Average (%)** |  | 59.66 |  | 32.42 |  | 7.92 |  | 100.00 |  | 4.58 |  | 78.33 |  | 22.86 |  | 29.00 |  | 64.72 |  | 4.12 |

### 3.4 Young males are more likely to suffer myocarditis and pericarditis following COVID-19 mRNA vaccination

To confirm the early evidence that young males were most susceptible to the adverse reaction of myocarditis and pericarditis (1), we compared spontaneous reporting and the literature up until 16 March 2022 to determine whether this signal has been maintained since initially identified. Of the myocarditis and pericarditis events reported to the Yellow Card scheme, 60.92% were from males with a trend towards increased frequency in younger age groups (Table 1; Figure 3A). This trend of more frequent reporting from males was similar across the three regions assessed, with 72.92% in the US and 60.75% in the EU/EEA (Figure 3A), as well as similar reporting trends between the vaccine types (Figure 3B).

**Figure 3:**
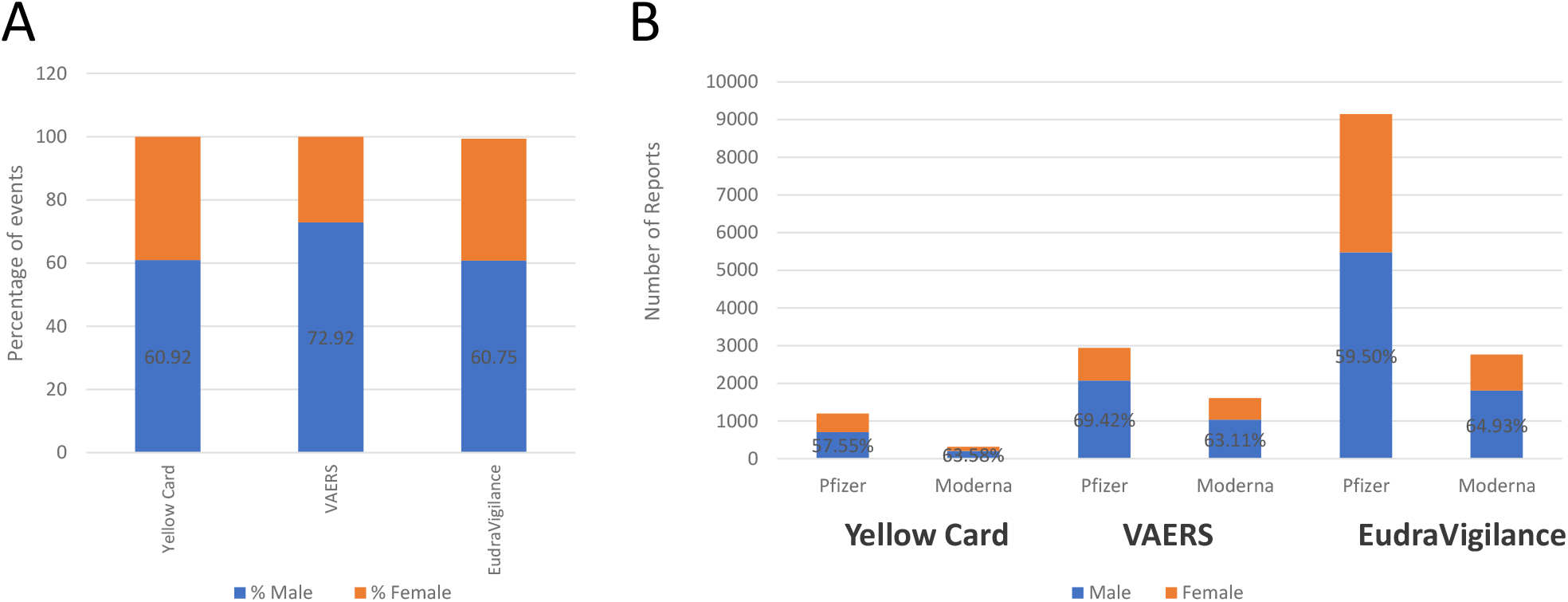
Myocarditis and pericarditis reports separated by gender. (A) Percentage of males and females experiencing myocarditis and pericarditis reported to each reporting system (Yellow Card, VAERS and EudraVigilance). Bars represent the total of all myocarditis and pericarditis reports to each system, separated by gender, where bars do not meet 100% indicates reports where gender was not specified. Values indicate the percentage of males experiencing these events. (B) Reports in each region were stratified by vaccine manufacturer as well as gender experiencing these events. Stacked bars represent the total number of reports, where gender was specified, from each region for each vaccine type. Values depict the percentage of males that experienced these events.

Most reports of myocarditis in the US were from vaccinees aged 18-39 years (n=1303, 47.3% of 2757 reports), while in the EU 79.2% (n=4431) of reports of myocarditis where from people aged 18-64 years (Table 4). In the US, 59.39% of pericarditis events were reported from males (Table 2) and the age distribution of those who reported pericarditis was wide, covering many age categories (Table 2). In the EU, 50.43% of the reported pericarditis events occurred in males (Table 4). The trend in age was less distinct, with children (3-11 years) and the elderly (aged older than 85 years) populations accounting for 35.7% and 43.97% of reported cases, respectively (Table 4).

Analysis of the literature determined that on average 60.31% of myocarditis and/or pericarditis events following COVID-19 mRNA vaccines occurred in males (range 50.00-100.00%). Results of these studies indicate that the incidence for myocarditis and pericarditis was higher for males than for females. Due to differences in study design it was not possible to determine the age group most susceptible to myocarditis and pericarditis from the literature as some studies were focused on adults only or children only meaning appropriate comparisons could not be carried out.

### 3.5 Most myocarditis and pericarditis events are reported following the second mRNA vaccine dose

Analysis of data following booster programme roll out has enabled detailed analysis into the frequency of reports following each dose of a COVID-19 mRNA vaccine. Data for the US is stratified by dose and demonstrates that the majority of reported myocarditis and pericarditis events occurred following the second dose (Figures 4A, 4B). For Comirnaty, of the total 1824 myocarditis events reported to VAERS, 956 (52.41%) followed the second dose, while 383 (21.00%) were reported after a single dose of the vaccine (Table 3a). Similarly, 45.78% of pericarditis events were reported to VAERS following two doses of Comirnaty (n=532 of 1162 reported events where vaccine dose was specified; Table 3a). For Spikevax, 384 of 933 (41.16%) reported events of myocarditis and 305 of the 707 (43.14%) reported events of pericarditis occurred following two doses of the vaccine (Table 3b). Approximately 10% of myocarditis and pericarditis events had been reported following a third dose for both vaccines up to 14 March 2022 (Tables 3a and 3b).

**Figure 4:**
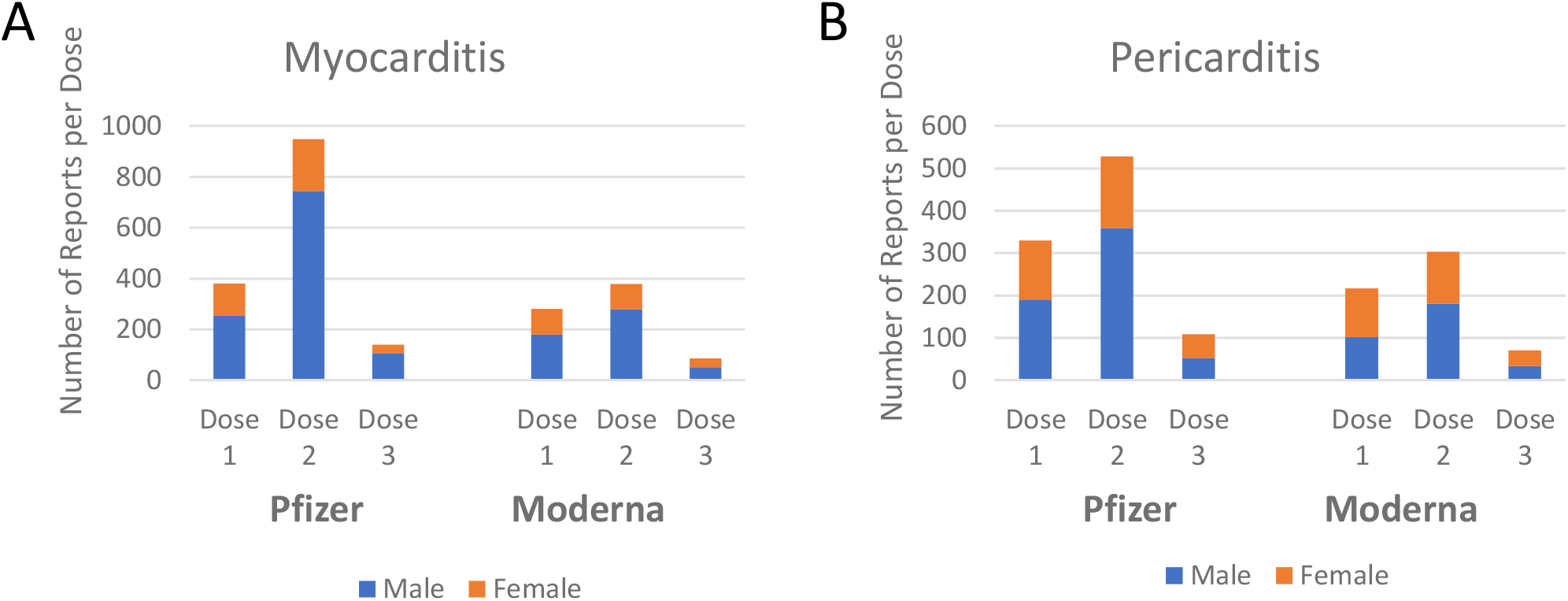
Myocarditis and pericarditis events following COVID-19 mRNA vaccination separated by dose. Data from the VAERS (US) reporting system was separated by reaction type; myocarditis (A) and pericarditis (B), vaccine manufacturer and by dose received. Stacked bars represent the male and female reports of each condition according to the dose received.

This data is in agreement with other studies where 64.72% (range 25.00% and 91.78%) of myocarditis and pericarditis events occurred following the second vaccine dose (Table 6).

## 4.0 Discussion

There have been a small number of reports of myocarditis and pericarditis following exposure to mRNA COVID-19 vaccines in each database examined, considering the number of people who have received a COVID-19 vaccine in each region. In all spontaneous reporting systems and for both mRNA vaccines, the reporting rate of myocarditis was higher than that of pericarditis. This may be true, or it may also reflect that the diagnosis of myocarditis is relatively more straightforward. In the UK and EU/EEA, reporting rates of myocarditis and pericarditis were higher following Spikevax compared with those for Comirnaty. However, this was not observed for the US. A full dose of Spikevax used for first and second doses contains 100 micrograms of mRNA nucleotides, whereas a full dose of Comirnaty contains 30 micrograms of the mRNA material (4, 5). This is one possible reason for the higher reporting rate of myocarditis and pericarditis observed for Spikevax in the UK population. A half-dose of Spikevax is used for booster vaccination doses; this still contains more genetic material than Comirnaty (4). Therefore, it will become increasingly important as more people receive third and subsequent booster doses of mRNA COVID-19 vaccines to monitor the frequency and severity of myocarditis and pericarditis following exposure to these vaccines.

The UK had the highest reporting rate for both myocarditis and pericarditis following mRNA vaccines, particularly for events following Spikevax. Reporting rates from the UK showed higher variability compared with those of the US and EU/EEA; reporting rates were higher for myocarditis and pericarditis following Spikevax in the UK, compared to EU/EEA and US reporting rates and incidence reported in the literature. This variability is potentially due to the fact that the UK MHRA Yellow Card scheme had the lowest number of spontaneous reports (Figure 1A) as well as the lowest vaccination coverage for Spikevax leading to uncertainty and variability. Analysis of multiple sources was thus essential to provide accurate reporting rates. It is possible that the UK and EU/EEA have stronger spontaneous reporting systems compared with the US, which may explain the higher reporting rates observed for this population. The frequency of events noted by regulators, the World Health Organisation, and in the vaccines’ summaries of product characteristics (SmPC) suggest that myocarditis and pericarditis are very rare, occurring in less than one in 10,000 vaccine doses administered (2, 5-7, 11). Our calculated reporting rates for myocarditis and pericarditis following mRNA vaccines in each of the UK, US, and EU/EEA were consistent with this. However, underreporting of the events to regulators is possible, therefore it may be that the events of myocarditis and pericarditis are ‘rare’ events (more frequent than 1/10,000) rather than ‘very rare’ (less frequent than 1/10,000) events as suggested. Conversely greater reporting may have occurred due to the public interest in adverse reactions linked with COVID-19 vaccines, thus analysis of multiple data sources was used here to overcome these potential limitations. In October 2021, the Pharmacovigilance Risk Assessment Committee (PRAC) of the European Medicines Agency announced a plan to review the risk of myocarditis and pericarditis following mRNA vaccines (29). It is therefore possible that updates to the vaccines’ SmPC will be required once this review is complete.

When examining the demographics of vaccinees who had reported myocarditis or pericarditis following mRNA vaccines, both events appear to more commonly affect males compared with females, particularly amongst vaccinees of younger age categories (Tables 2-4). This is consistent with early reports surrounding these events, where it was suggested that younger males appear at higher risk of myocarditis and pericarditis following mRNA vaccines (1). Demographic data was available for VAERS and EudraVigilance populations separated by myocarditis and pericarditis; in these databases results were similar with more than 70% of myocarditis and more than 50% of pericarditis events reported in males (Tables 2 and 4). These results are consistent with results of pharmacoepidemiological studies in the published literature; in the studies where sex was reported, more than 60% of patients were male (Table 5) (30-35). This is an interesting finding, as it has been previously suggested that over 70% of reports to VAERS involve females (31). It is possible that the event was missed or misclassification occurred in older adults, particularly in older individuals in the three populations of interest who were vaccinated prior to the signal emerging. It is also possible that some of the symptoms of myocarditis and pericarditis, for example chest pain and breathlessness, were attributed to other cardio-respiratory conditions in older people. Nonetheless, it is known that most cases of myocarditis (any cause) occur in young adults, with males more commonly affected than females; this supports the results observed in this study (36, 37). Alternatively, vaccine roll-out in each of the regions may have affected the results observed in some of the countries, including availability of different vaccines and corresponding age distributions of those receiving each vaccine in different regions; for instance in the UK, mRNA vaccines were more frequently used in younger age groups, while older vaccinees may have been more likely to receive an adenovirus vector vaccine.

We appreciate that there will be regional differences in COVID-19 strains as well as endemic viruses circulating in the populations, and these may have been the underlying cause of myocarditis and pericarditis in the reporting populations examined. However, it is not possible to identify or quantify these in spontaneously reported data. In order to overcome these differences, we have analysed data from three different spontaneous reporting systems and worldwide data from the literature to identify trends in this very rare event. As this is a very rare event with small numbers of people affected, it is important to bring together data from around the world to identify trends that may not be seen within one population. Further analysis is required in discrete populations, if appropriate, to better stratify patients, aiding classification of groups according to risk factors for adverse events following vaccination.

Data on vaccine dose were only available from the VAERS database. Most cases reported to VAERS followed a second dose of vaccine (Tables 3a and 3b). This is consistent with the early signal which emerged in Israel, where 121 of the 148 reported cases of myocarditis occurred around the time of the second dose of COVID-19 vaccine (1). Furthermore, similar results were observed by Hajjo et al., who found only a very small number of events occurring after a third dose in their analysis of VAERS data (38).

Pharmacoepidemiological studies identified by systematic review (Table 5) were consistent with results found in spontaneously data from VAERS and EudraVigilance (Tables 2-5). Young males more frequently reported myocarditis in each of these studies and found in our analysis of spontaneous reporting data. Furthermore, reports were more frequent following a second dose of mRNA vaccine, although time intervals between doses was not specified and may also differ between regions. Therefore incorporation of the evidence on dosing effects from the VAERS system and the literature indicated that following the second dose was when the most myocarditis and pericarditis events occurred. Continuous monitoring of this will be required, especially following subsequent doses of mRNA vaccines.

Myocarditis has been observed historically following vaccination, including after smallpox, influenza, and hepatitis B vaccines; prior to COVID-19 0.1% of reports to VAERS between 1990 and 2018 had been in relation to myopericarditis (13). Furthermore, myocarditis is known to occur after a range of viral infections, including coronaviruses which cause Middle East Respiratory Syndrome (MERS) and COVID-19, with viral infection the most common cause of myocarditis (39-41). This study provides evidence that younger vaccinees more frequently report myocarditis and pericarditis following mRNA COVID-19 vaccines compared with older vaccinees, and reports are more frequent following the second dose. Results were consistent between each of the three data sources used. This is an important finding, because as vaccination programmes around the world progress, rates of myocarditis and pericarditis are likely to increase. The effect of booster vaccinations with mRNA COVID-19 vaccines on the development of myocarditis and pericarditis is largely unknown. Furthermore, mRNA COVID-19 vaccines (particularly Spikevax) are being supplied to the COVAX initiative for distribution throughout low- and middle-income countries, where diagnostic imaging and access to healthcare is more difficult (42, 43). Regulatory authorities should continue to monitor the effects that mRNA vaccination might have on the heart in the populations for which they are responsible. The proportions of young people are higher in low- and middle-income countries’ populations compared to high income countries. Issuing diagnostic criteria and treatment protocols for myocarditis and pericarditis with mRNA COVID-19 vaccines that take into consideration the capabilities of the local healthcare system are also important.

### 4.1 Limitations

Vaccination policies in the three regions may have biased the results towards a higher number of adverse events reports myocarditis and pericarditis from younger vaccinees compared with older vaccinees. In each of these regions, younger people were more likely to have received mRNA vaccines, which may have contributed to higher reporting rates of myocarditis and pericarditis in younger vaccinees. The frequency of reported events per age group was presented as a crude number, and reporting rates could only be calculated as an overall estimate rather than stratified by age; based on the data available for vaccinations administered, it was not possible to determine the proportion of all vaccinees per age group who reported an event of myocarditis or pericarditis. This is very important, because it is likely that the reporting rate of myocarditis and pericarditis with mRNA COVID-19 vaccines will be higher in young people and even higher in young men if the reporting rates are stratified by age and sex. The regulatory authorities and Marketing Authorisations Holders (MAHs) need to follow up reports of these conditions with reporters to obtain as much information and make this information available publicly. Myocarditis and pericarditis following mRNA COVID-19 vaccines is an area which requires further research.

The data sources for this study were spontaneous reporting systems of the UK, US, and EEA. All spontaneous reporting systems have well-known limitations including missing information, and reporting bias caused by publicity surrounding a particular adverse event (44). Misclassification of myocarditis and pericarditis, or differing definitions of these events between the regions analysed, is also possible particularly before these events attracted publicity or among older age groups. The definition and diagnosis of “Myocarditis” and “Pericarditis” are based on the reporters’ statements, and are not usually validated by the regulatory authority receiving the report. Under-reporting is a major limitation of spontaneous reporting; even with the intense publicity and global attention on COVID-19 vaccine safety, it is possible that not all cases are reported to regulatory authorities (45, 46). Furthermore, a report to spontaneous reporting systems indicates suspicion that the event was associated with the vaccine, it does not confirm that the vaccine caused the event (11, 44). Further assessment is required to determine causality for each report. Finally, it is not possible to estimate incidence rates using spontaneous reports, and there is no unvaccinated comparison group (44).

Using publicly available data introduced some challenges, as the level of detail available was limited and varied between data sources. Data on the vaccine dose on which the reported events of myocarditis and pericarditis occurred were only available for the US VAERS population. Information contained within individual reports is not routinely made available, however these comprise important clinical information that would allow better understanding of each case. Such details should be made publicly available. Better transparency is needed to allow more robust research using spontaneous reporting to be undertaken.

Due to the limited published research into myocarditis and pericarditis following mRNA COVID-19 vaccines, we included all pharmacoepidemiological study designs (except case reports and case series) and considered all study populations and all study periods for inclusion. The purpose of systematically reviewing the literature was to determine whether results from our analyses of spontaneous reports were consistent with other evidence currently available. Due to the short time period since vaccine programmes were initiated and the heterogenous nature (regional differences, age, sex, and disease status of participants) of the included studies, no pooling of data, syntheses, or meta-analyses could be completed. It should be noted that there were no formal assessments of publication bias during the systematic literature review. However, a CASP checklist was completed for each included study, which deemed the research to be of sufficient quality for inclusion. Nonetheless, each study had limitations which should be considered when interpreting their results. The possibility of publication bias was not formally analysed.

Further pharmacoepidemiological studies are urgently needed to address many of the limitations of spontaneous reporting in understanding myocarditis and pericarditis following mRNA COVID-19 vaccines including more accurate estimates of the frequency, better understanding of the clinical course and the effects of these events on quality of life. It is also important to compare the incidence and characteristics of these events with recipients of other non-mRNA COVID-19 vaccines and unvaccinated people. However, these studies will take time to be conducted.

## 5.0 Conclusions

This study adds to existing evidence that younger vaccinees more frequently report myocarditis and pericarditis following mRNA COVID-19 vaccines compared with older vaccinees, and reports are more frequent following the second dose. These events are very rare or possibly rare according to the estimated reporting rates from spontaneous adverse reactions. The events were more frequently reported amongst males, and most reports came from vaccinees aged under 30 years. The clinical course of these events is typically mild, with full recovery in most cases.

The study brings together spontaneously reported adverse event data from three regions. Consistencies in the reporting rates and trends of myocarditis and pericarditis within the three data sources utilised suggest that results may be generalisable to other populations in which mRNA vaccines are used. However, limitations of the data sources used and biases which may have affected results should be considered. It is important that regulatory authorities continue to monitor the effects of mRNA vaccines on the heart, particularly as vaccine programmes progress to include younger vaccinees in many parts of the world. Myocarditis and pericarditis following mRNA COVID-19 vaccines is an area which requires further research, especially in children and adolescents and following third and subsequent (booster) doses. Pharmacoepidemiological studies are urgently needed to address many of the limitations of spontaneous reporting in understanding myocarditis and pericarditis following mRNA COVID-19 vaccines including more accurate estimates of frequency, a better understanding of the clinical course, and the effects on quality of life. However, they will take time to be conducted. We believe that the data we describe here will enhance the understanding of these conditions and help with identification of potential sources of mechanisms of vaccine-associated myocarditis and pericarditis by identifying which populations are most likely to suffer these adverse events following COVID-19 mRNA vaccination.

## Data Availability

CASP checklists for assessing quality of each study included in systematic review are available on request.

## Declarations

### Transparency statement

The manuscript’s guarantor affirms that the manuscript is an honest, accurate, and transparent account of the study being reported, and that no important aspects of the study have been omitted.

### Ethics approval

Ethics approval was not required.

### Funding

No external funding was received for the preparation of this manuscript.

### Conflicts of interest

All authors have completed the Unified Competing Interest form (available on request from the corresponding author) and declare: The Drug Safety Research Unit (DSRU) is a registered independent charity (No. 327206) associated with the University of Portsmouth. The DSRU receives donations and grants from pharmaceutical companies; however, the companies have no control over the conduct or publication of its studies. The DSRU has received grants to conduct unconditional studies on the Oxford/AstraZeneca COVID-19 vaccine and is in negotiations to receiving grants for conducting CPRD studies for Pfizer, Moderna, and Janssen COVID-19 vaccines. The DSRU has conducted benefit-risk studies on products for COVID-19, including remdesivir, lopinavir/ritonavir, chloroquine and hydroxychloroquine, and convalescent plasma. Professor Shakir is the principal investigator for an active surveillance study for the Oxford/AstraZeneca vaccine, but this assessment is unrelated to this study. Professor Shakir has been a member of Data Safety Monitoring Boards for Ipsen, Biogen, and Diurnal. None of these companies have any involvement with COVID-19 vaccines. Professor Shakir was invited by AstraZeneca to advise on the events of thrombosis with thrombocytopenia with the COVID-19 vaccine and to be a member of an advisory committee on a safety study of the Oxford/AstraZeneca vaccine in Europe. Samantha Lane and Alison Yeomans have no conflicts of interest with regard to this study.

### Authors’ contributions

SL and AY were responsible for data acquisition, analyses, and interpretation. All authors were responsible for study conception, drafting and reviewing the manuscript, and approval of the final version for publication.

## Notes

### Author Declarations

Ethics approval was not required.

### Summary of Updates

Updated throughout following peer-review. More recent data added. Systematic review has been re-run and more studies included. Paper re-structured. Figure 1 added. All tables and figures revised. Table 6 added. Conclusions remain unchanged.

